# Systematic Review and Meta-Analysis of the Effect of Adverse Childhood Experiences (ACEs) on Brain-Derived Neurotrophic Factor (BDNF) Levels

**DOI:** 10.1101/2022.08.16.22278345

**Authors:** Neha Vyas, Courtney E. Wimberly, M. Makenzie Beaman, Samantha J. Kaplan, Line J.H. Rasmussen, Jasmin Wertz, Elizabeth J. Gifford, Kyle M. Walsh

**Affiliations:** Duke University, Trinity College of Arts and Sciences, Durham, NC, USA; Duke University School of Medicine, Durham, NC, USA; Duke University Department of Neurosurgery, Durham, NC, USA; Duke Children’s Health and Discovery Initiative, Durham, NC, USA; Duke University Department of Psychology and Neuroscience, Durham, NC, USA; Department of Clinical Research, Copenhagen University Hospital Amager and Hvidovre, Hvidovre, Denmark; University of Edinburgh, Department of Psychology, Edinburgh, UK; Duke University Sanford School of Public Policy, Center for Child and Family Policy, Durham, NC, USA

**Author notes:** <u>Corresponding author:</u> Kyle M. Walsh, DUMC Box 3050, Durham, NC 27710.

**Keywords:** Brain-Derived Neurotrophic Factor, Adverse Childhood Experiences, Biomarkers, Systematic Review, Meta-Analysis

## Abstract

There is continued interest in identifying dysregulated biomarkers that mediate associations between adverse childhood experiences (ACEs) and negative long-term health outcomes. However, little is known regarding how ACE exposure modulates neural biomarkers to influence poorer mental health outcomes in ACE-exposed children. To address this, we performed a systematic review and meta-analysis of the impact of ACE exposure on Brain Derived Neurotrophic Factor (BDNF) levels - a neural biomarker associated with the onset of mood disorders. Twenty-two studies were selected for inclusion within the systematic review, ten of which were included in meta-analysis. Most included studies retrospectively assessed impacts of childhood maltreatment in clinical populations. Sample size, BDNF protein levels in ACE-exposed and unexposed subjects, and standard deviations were extracted from ten publications to estimate the BDNF ratio of means (ROM) across exposure categories. Overall, no significant difference was found in BDNF protein levels between ACE-exposed and unexposed groups (ROM: 1.05; 95% CI: 0.91-1.22). Non-significant elevation of BDNF levels in ACE-exposed subjects was observed among studies specifically investigating childhood maltreatment and among studies measuring BDNF in serum, but these analyses were limited by high between-study heterogeneity. Studies that measured BDNF levels in subjects prior to age 20 revealed elevated levels in the ACE-exposed group, which was not observed in studies measuring BDNF levels later in life. These results support continued investigation into the impact of ACE exposure on neural biomarkers and highlight the potential importance of analyte type and timing of sample collection on study results.

## Introduction

The term adverse childhood experiences (ACEs) refers to potentially traumatic events occurring during childhood (*e.g*., physical abuse, emotional abuse, sexual abuse) (Felitti et al., 1998). In 2019, a study by the Centers for Disease Control and Prevention (CDC) reported 61% of individuals within the United States had been exposed to at least one traumatic incident during childhood (Merrick et al., 2019). Exposure to ACEs has been found to confer enduring negative impacts on the physical and mental health of ACE exposed children (Hughes et al., 2017; Kalmakis & Chandler, 2015). Due to the prevalence of ACE exposure and its impact on a diverse set of physiological systems, a field of research has emerged that focuses on identifying quantifiable biomarkers of ACE exposures. Such biomarkers could elucidate the biological pathways through which ACEs negatively impact health and could be further leveraged to investigate mechanisms for enhancing physical and psychological resilience. Prior research has focused primarily on dysregulation of the endocrine system and the hypothalamic-pituitary-adrenal (HPA) axis, primarily by investigating the effect of ACEs on cortisol levels (K. S. Dempster, O’Leary, MacNeil, Hodges, & Wade, 2021) and inflammatory biomarkers (Baumeister, Akhtar, Ciufolini, Pariante, & Mondelli, 2016; Kuhlman, Horn, Chiang, & Bower, 2020). Substantially less is known regarding the effect of ACEs on the developing nervous system, despite suggestive evidence of ACE-driven, detrimental impacts on the long-term mental health of ACE exposed children during adulthood (Schauss et al., 2019).

ACEs are a major contributing factor to early mortality, and exposure to even a single ACE has been found to increase risk of chronic diseases, such as heart, lung, and liver disease, in addition to cancer, type 2 diabetes, obesity, and psychiatric disorders (Deighton, Neville, Pusch, & Dobson, 2018; Godoy et al., 2021; Grummitt et al., 2021), but how ACE exposure directly perturbs diverse physiological systems remains poorly defined (Deighton et al., 2018; Kalmakis & Chandler, 2015; Petruccelli, Davis, & Berman, 2019). ACE-associated increases in inflammatory markers, including C-reactive protein (CRP), Tumor Necrosis Factor-α (TNF-α), and Interleukin-6 (IL-6), have been observed to partially mediate subsequent development of obesity and other chronic conditions (Kylie S Dempster, O’Leary, MacNeil, Hodges, & Wade, 2020). ACE-mediated alterations in the HPA axis have also been associated with dysregulated cortisol levels in children, which can subsequently affect biomarkers and functioning of other biological systems (Deighton et al., 2018). Increased exposure to ACEs is associated with detrimental impacts on cardiometabolic health, such as increases in blood pressure, body mass index (BMI), waist circumference, and triglyceride levels, all of which elevate cardiovascular disease risk (Godoy et al., 2021; Pretty, D O’Leary, Cairney, & Wade, 2013). Thus, it is evident that exposure to ACEs affects physical health in a manner that spans several physiological systems.

Beyond the multifaceted effects of ACEs on one’s physical health, ACE exposure also has strong associations with psychiatric diagnoses later in life, such as depression, PTSD, anxiety, and substance abuse disorders (Deighton et al., 2018; Kalmakis & Chandler, 2015). Alterations in neural development due to ACE exposure may underlie this link, but the mechanisms mediating such a relationship remain poorly defined (Schauss et al., 2019; Wilson & Perez Vallejos, 2021). Childhood, especially early childhood, is a time of extensive neural development, as children begin to accumulate memories and strengthen the neuronal connections that underlie memory formation (Buss et al., 2007). Exposure to ACEs at this stage of life may impair proper memory consolidation and increase a child’s likelihood for developing phobias, anxiety disorders, or other trauma-related disorders (Schauss et al., 2019). Additionally, childhood is also a time of extensive brain growth, and traumatic experiences during childhood have been observed to impair growth of specific brain regions. For instance, Navalta, McGee, and Underwood (2018) found that childhood exposure to traumatic experiences was associated with decreased volumes of the hippocampus, amygdala, medial prefrontal cortex, and limbic structures in adulthood. Because these structures are each essential for regulation of emotional arousal, developmental perturbation of any of these structures could increase the likelihood of developing anxiety disorders or mood dysregulation (Navalta et al., 2018). Thus, the potential impact of ACEs on the structure of the nervous system may explain an increased propensity toward mood disorders.

One biomarker hypothesized to mediate ACE driven impacts on nervous system function is brain derived neurotrophic factor (BDNF). A member of the neurotrophin family of growth factor proteins, BDNF binds to Tropomyosin receptor kinase B (TrkB) receptors on neurons of the central and peripheral nervous systems where it supports neuronal survival and encourages neurogenesis (Binder & Scharfman, 2004). BDNF has predominantly been treated as a biomarker of synaptic plasticity (Benedetti et al., 2017; Kim, Watt, Ceballos, & Sharma, 2019), and due to its secretion by activated lymphocytes and the ability of IL-6 and TNF-α to augment BDNF release from the nervous system (Jin, Sun, Yang, Cui, & Xu, 2019; Kraneveld et al., 2014), BDNF activity resides at the intersection of neural development, memory formation, and neuroimmunology (Bekinschtein, Cammarota, Izquierdo, & Medina, 2007; Calabrese et al., 2014; Kim et al., 2019). In addition to BDNF’s modulation of the immune axis, it has also been implicated in instigating dysregulation of the HPA axis, with elevated BDNF levels contributing to hyperactivity of the HPA axis – a common molecular change seen in ACE-exposed children (K. S. Dempster et al., 2021; Naert, Ixart, Maurice, Tapia-Arancibia, & Givalois, 2011). Furthermore, because BDNF can be non-invasively measured in saliva and plasma, it is frequently investigated as a potential biomarker of ACE exposure – one that is hypothesized to mediate the contributions of ACEs to nervous system dysfunction and the development of neuropsychiatric illnesses. BDNF levels have been implicated in the development of mood disorders which have been associated with ACE exposure, such as bipolar disorder, depression, and schizophrenia (Bocchio-Chiavetto et al., 2010; Gaglia, 2021; Kauer-Sant’Anna et al., 2007).

While BDNF has been frequently investigated, findings have predominantly explored the impact of ACE exposure on methylation of the *BDNF* gene promoter regions, and subsequent effects on the expression of the BDNF gene, both in blood and in neural tissues (Jiang, Postovit, Cattaneo, Binder, & Aitchison, 2019; Neves, Dinis-Oliveira, & Magalhães, 2019; Pilkay, Combs-Orme, Tylavsky, Bush, & Smith, 2020). However, findings have remained largely inconclusive due to the key drawback of using methylation-based studies of BDNF, as DNA methylation varies in association with age, sex, and ethnic of subject, as well as the tissue type from which DNA is extracted (Tsai, 2018). While methylation-based studies can help reveal the functional genetic mechanisms underlying variability in BDNF gene expression, directly measuring BDNF protein level and its association with prior ACE exposure can substantially simplify interpretation of results, particularly when BDNF in protein is measured from a single, consistent type of biospecimen.

Thus, we sought to clarify the relationship regarding the impact of ACE exposure and BDNF levels via specifically analyzing protein levels, which can be more reliably quantified and thus will enable generalizability of results. We conducted a systematic literature review and meta-analysis to identify and integrate data from all published studies investigating the impact of ACE exposure on BDNF protein levels. We further performed diagnostics to evaluate the quality and consistency of the published literature, and subgroup analyses to explore potential heterogeneity due to difference in study designs and the specific types of ACEs evaluated. By rigorously amalgamating data across studies, our results can help to clarify the relationship between ACE exposure and BDNF levels, reveal current knowledge gaps, and inform future mechanistic research into how ACEs may impact long-term neurological health and disease.

## Methods

### Definition of ACEs

ACEs were first defined in the 1998 CDC-Kaiser Study (Felitti et al., 1998), which delineated specific traumatic exposures during childhood that were found to have long-term, detrimental impacts on the child’s physiological and mental health. We employed this definition of ACEs, including ten distinct exposures: physical abuse, emotional abuse, sexual abuse, physical and emotional neglect (including extreme poverty/deprivation), presence of parent with mental illness, incarcerated family member, exposure to domestic abuse or intimate partner violence, presence of parent with history of substance abuse, and family separation (parental death, foster care, or parental divorce).

### Search Strategy

A medical librarian (SK) with expertise in the design and conduct of systematic literature reviews assisted with development of a search strategy that utilized a mix of keywords and subject headings representing the concepts of ACEs (*e.g*., child abuse, domestic violence, family separation, poverty), biomarkers (*e.g*., endocrine, immune, cardiovascular, and nervous), stress reaction, and children. The search underwent internal peer-review by another medical librarian with similar expertise. A full accounting of search terms and strategies appears in **Supplementary Table 1**. The search study was applied to four databases: MEDLINE via Ovid Wolters Kluwer, Embase via Elsevier, Scopus via Elsevier, and APA PsycINFO via EBSCO and was conducted on April 27, 2020. When possible, conference abstracts and non-human studies were removed. Results were compiled into EndNote, imported into Covidence for screening, and duplicate studies were removed.

### Study Screening and Selection

Title and abstract screening were completed by all authors. Abstract screening was conducted by a minimum of two authors, of which one author must have been a senior team lead (EG, KW, LR, and JW). Criteria for inclusion of abstracts required mention of biomarkers, ACEs, or exposure to childhood trauma within the sample. Screening discordances were resolved through discussion with the full team. Full-text manuscripts of selected abstracts were then uploaded to REDCap for full-text screening. A sample REDCap screening tool can be found in **Supplementary Figure 1**.

Selected studies were required to 1) report both ACE exposure and measurement of relevant biomarkers, and 2) quantify the impact of ACEs on selected biomarker(s). Studies were excluded if they: 1) did not mention both ACE exposure and the biomarker measured, 2) were conducted in animal models, 3) were not original research (reviews, editorials, or commentaries), 4) were reports of single patients (*i.e*., case-reports), and 5) were unavailable in English. Following identification of all studies that passed the full-text inclusion/exclusion criteria, all studies that were annotated as quantifying neurotransmitter levels were individually evaluated to retain only those studies which specifically measured BDNF protein levels. The citations list of all included studies was also screened using the original inclusion/exclusion criteria to identify any additional relevant studies not captured by the search strategy. For studies that quantified BDNF protein levels but did not report specific numeric levels in the manuscript, we contacted study authors in order to obtain these data (*n=13*). If no response was received, the study was excluded from the meta-analysis (*n=11*)

### Data Extraction + Synthesis

All publications retained following full-text screening were uploaded into REDCap for preliminary data extraction. Data extracted from publications included: sample description, sample size, specific ACE exposure and time of exposure, specific biomarker and time of measurement, study method, and summary of findings. Exclusion reason was also cited for full-text publications that were reviewed but did not meet inclusion criteria. Following extraction of these preliminary data, all studies with mention of neurotransmitters were isolated and studies analyzing BDNF protein levels were selected for inclusion in the systematic review. All studies measuring the relationship between ACEs and BDNF levels were reviewed, and the following data was extracted: sample size and study population, type of ACE measured and timing of measurement (*e.g*., during childhood, retrospectively in adulthood), BDNF measurement methodology and timing, and general impact of ACE exposures on BDNF levels.

From these studies, a small subset of studies was isolated for inclusion in the meta-analysis, all of which explicitly quantified mean BDNF protein levels in an ACE-exposed sample and an unexposed comparator population. Data extracted from studies included within the meta-analysis was: ACE exposure(s), BDNF measurement methodology, sample characteristics, mean BDNF levels (in ACE-exposed and unexposed), and standard deviation of BDNF measures (in ACE-exposed and unexposed). Mean BDNF levels and standard deviations were used to calculate the ratio of means (ROM) for ACE-exposed versus ACE-unexposed subjects, including 95% confidence intervals. Fixed and random-effects meta-analysis of the ROM across studies was conducted in R-Studio using the meta, metafor, and dmetar packages to calculate a summary ROM with its 95% confidence interval, study weight (based on sample size), between-study heterogeneity via the *I*^2^ statistic, and to assess the presence of publication bias using the Egger’s regression asymmetry test, and to generate forest plots. Mixed-effects meta-analysis was also performed in R-Studio to explore heterogeneity in the summary-level ROM across strata of study design.

## Results

### Search Strategy Results

Following application of the search strategy to databases, 11,845 abstracts were uploaded onto COVIDENCE. 3,710 of these entries were excluded as they were duplicates, and an additional 7,009 records were excluded following abstract and title review. 1,126 articles were selected for full-text screening, of which 145 were excluded due to lack of adherence with inclusion criteria. Of the 981 studies screened in, thirty-nine full-text articles were recorded as measuring neurotransmitters, including BDNF, as a biomarker of ACE exposure and were rigorously evaluated for potential inclusion in our meta-analyses. Seventeen studies were excluded because they did not measure BDNF protein levels (9 measured different neurotransmitters and 8 measured DNA methylation levels at the *BDNF* gene locus). Twenty-two studies were thus included in our systematic review, of which ten quantified reported mean BDNF levels and standard deviations in ACE-exposed and unexposed subjects (or study authors provided these data via personal communication) and were included in the meta-analysis. A detailed overview of the flow of studies through the screening process is presented in a PRISMA diagram (**Figure 1**).

**Figure 1.**
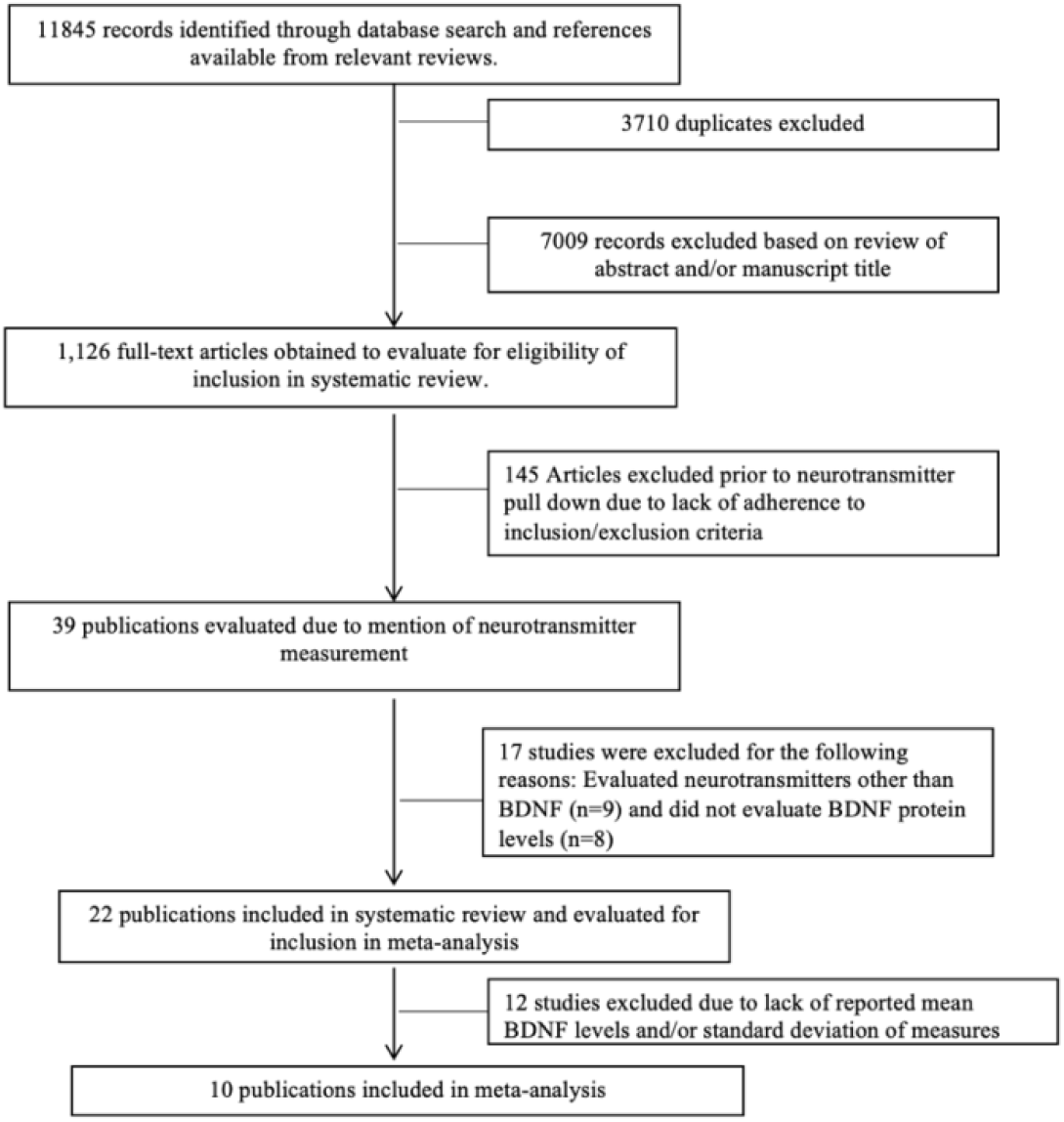
PRISMA flowchart depicting study selection strategy and reasons for exclusion.

### Key Findings from Systematic Literature Review

Of the twenty-two studies included within the systematic review, eighteen studies explored the impact of ACEs on clinical populations, and four studies had a sample representative of the general population. The majority of studies (*n=16*) investigated childhood maltreatment as the ACE exposure. Specifically, sexual abuse (n=*15*), physical abuse (*n=12*), emotional abuse (*n=9*), and neglect (*n=10*) were all evaluated within this subset of studies. Other ACE exposures measured include: parental mental health (*n=3)*, parental substance abuse (*n=2)*, parental incarceration (*n=1*), parental death (*n=1*), peer victimization and bullying (*n=1*), family dysfunction (*n=1*), and overall childhood trauma (*n=1*). ACE exposure was predominantly measured retrospectively in adulthood (*n=15*) rather than during childhood while exposure was ongoing (*n=7*).

BDNF protein levels were primarily measured from serum (*n= 14)*, plasma (*n= 5)*, and whole blood (*n=3)*. Levels were measured predominantly with an ELISA assay (*n=21*); however, one study utilized a Luminex assay (*n =1*). BDNF measurements were conducted both retrospectively in adulthood (*n=15)* and during childhood while the ACE was ongoing (*n=7)*. Qualitatively, nine studies reported elevated BDNF levels in the ACE-exposed group versus the ACE-unexposed group, while eleven studies reported lower BDNF levels in the ACE-exposed group, and two studies reported no detectable difference in BDNF levels between exposure groups. Of the four studies measuring ACE exposure and BDNF levels in a representative sample drawn from the general population, two studies reported higher BDNF levels in the ACE-exposed group, and two studies reported lower BDNF levels in the ACE-exposed group. For studies investigating clinical populations, seven studies reported higher BDNF levels in the ACE-exposed group, nine studies reported lower BDNF levels in the ACE-exposed group, and two studies found no detectable difference in BDNF levels across exposure groups. Detailed study characteristics appear in **Supplementary Table 2**.

### Key Findings from Meta-Analyses

Within the subset of studies included in the meta-analysis, six studies investigated the impact of ACEs on BDNF levels within clinical patients and four studies investigated subjects recruited from the general population. Childhood maltreatment was the most common ACE exposure (*n=6)* reported by studies. Within these six studies, exposure to sexual abuse (*n=6)*, physical abuse (*n=5)*, emotional abuse (*n=4*), and neglect (*n=3)* were specifically reported. ACE exposure was predominantly measured retrospectively in adulthood (*n=6)*. BDNF levels were most commonly measured in serum (*n=4)*, whole blood (*n=3)*, and plasma (*n=3)*. Six studies observed lower BDNF levels in the ACE-exposed group versus the unexposed group, and four studies observed elevated BDNF levels in the ACE-exposed group. Among studies looking at the general population, three studies reported lower BDNF levels in the ACE-exposed group, and one study reported higher BDNF levels in the ACE-exposed group. Three studies with clinic-based participants reported lower BDNF levels in the ACE-exposed group, and three studies reported higher BDNF levels in the ACE-exposed group. Detailed characteristics of studies included in the meta-analysis appear in **Table 1**.

**Table 1:** Characteristics of Studies Included in Meta-Analysis.

| Title of Study | ACEs Measured | BDNF Measurement Methodology | Comparator Sample (Sample Size) | ACE- Sample Mean BDNF Levels (Standard Deviation) | ACE+ Sample (Sample Size) | ACE+ Sample Mean BDNF Levels (Standard Deviation) | Mean Difference (ACE-minus ACE+) and 95% Normalized CI |
| --- | --- | --- | --- | --- | --- | --- | --- |
| Bortoluzzi et al. (2014) | Physical Neglect | Serum BDNF levels were measured with the use of an ELISA assay. | Adolescents with anxiety and exposure to low levels of trauma (*n*=50) | 37.87 ng/mL (13.08) | Adolescents with anxiety and exposure to high levels of trauma (*n*=10) | 54.05 ng/mL (20.21) | -16.05 ng/mL (-0.656, -0.198) |
| Bücker et al. (2015) | Childhood abuse (sexual, physical, and/or emotional) and neglect (physical and/or emotional) | Plasma BDNF levels were measured using the Sandwich ELISA method. | Healthy Controls (*n*=26) | 0.89 pg/mL (0.57) | Children in foster care /protective services exposed to measured ACEs prior to the age of 4 (*n*= 36) | 1.42 pg/mL (0.25) | -0.53 pg/mL (-0.858, -0.333) |
| do Prado et al. (2017) | Childhood Abuse (sexual, physical, and/or emotional abuse) and neglect (emotional and/or physical) | Plasma BDNF levels were measured with the use of an ELISA assay | Healthy Controls (*n*=27) | 347.96 pg/mL (224.11) | Adolescents with exposure to early life stress (*n*=30) | 200.07 pg/mL (97.48) | 147.9 pg/mL (0.162, 0.688) |
| Hauck et al. (2010) | Remote Trauma (not explicitly defined) | Serum BDNF levels measured using ELISA assay | Healthy Controls | 0.25 pg/ug (0.14) | PTSD patients with remote exposure to trauma | 0.39 pg/ug (0.16) | -0.14 pg/ug (-0.96,-0.16) |
| Sharma, Graham, Rohde, and Ceballos (2017) | Parental substance abuse | Serum BDNF levels measured using ELISA assays | Healthy social drinkers without family history of alcohol abuse | 424.96 pg/mL (324.89) | Healthy social drinkers without family history of alcohol abuse | 423.99 pg/mL (260.18) | 0.97 pg/mL (-0.363, 0.367) |
| Simsek, Uysal, Kaplan, Yuksel, and Aktas (2015) | Sexual abuse | Blood BDNF levels measured using an ELISA assay | Children with exposure to sexual abuse without PTSD (n=28) | 2.4 ng/mL (2.3) | Children with exposure to sexual abuse with PTSD (n=27) | 2.3 ng/mL (2.2) | 0.1 ng/mL (-0.454, 0.537) |
| Sordi et al. (2019) | Childhood abuse (sexual, physical, and/or emotional) | Blood BDNF levels were measured using the Sandwich ELISA method | Adults defined to have low trauma exposure (n=11) | 29.28 pg/mL (11.62) | Adults defined to have high trauma exposure (n=11) | 30.77 pg/mL (8.13) | -1.41 pg/mL (-0.337, 0.235) |
| Theleritis et al. (2014) | Childhood abuse (sexual and/or physical) and parental death | Plasma BDNF levels were measured using an ELISA assay | Unaffected Controls (n=152) | 25.845.0 pg/mL (6.4408) | First Episode Psychosis Patients having been exposed to physical and/or sexual abuse (n=95) | 24.7541. pg/mL (6.3805) | 1.09 pg/mL; (-0.021,0.106) |
| Trajkovska et al. (2008) | Parental mental health and parental death, | Blood BDNF levels measured using ELISA assay | Females at high risk for MDD diagnosis with 0-2 stressful life events | 19.2 ng/mL (4.8) | Females at high risk for MDD diagnosis with 3+ stressful life events | 18.5 ng/mL (4.1) | 3.1 ng/mL (0.296, 5.90) |
| Watt, Ceballos, Kim, Pan, and Sharma (2020) | Parental mental health, parental substance abuse, parental incarceration, and exposure to intimate partner violence | Serum BDNF levels were measured using an ELISA assay | ACE score less than or equal to 3 ($n = 30$) | 421.08 pg/mL (153.58) | ACE score greater than or equal to 4 ($n = 63$) | 377.34 pg/mL (132.07) | 43.74 pg/mL; (-0.0479, 0.257) |

Aggregate analysis was performed on ten studies from which mean BDNF levels and standard deviations for both the ACE-exposed and an unexposed comparator group could be ascertained. A random-effects meta-analysis of the ROM in ACE-exposed versus unexposed subjects was not significantly different from 1.0 (ROM = 1.05; 95% CI: 0.91-1.22) (**Figure 2**). While the meta-analytic ROM estimate was not significant, four studies reported significant differences in BDNF protein levels between the ACE-exposed and unexposed groups. Do Prad et al. reported significantly lower BDNF levels in the exposed group than the unexposed group (ROM = 0.57, 95% CI: 0.43-0.78). Bücker et al. (ROM = 1.60, 95% CI: 1.24-2.05), Hauck et al. (ROM = 1.56, 95% CI: 1.17-2.09), and Bortoluzzi et al. (ROM = 1.43, 95% CI: 1.11-1.83) reported significantly higher BDNF levels in the exposed group than the unexposed group. High between-study heterogeneity was observed (I^2^ = 0.81), and Cochran’s Q value also revealed statistically significant heterogeneity (p<0.01). Egger’s regression test indicated that no single study unduly influenced the results of the meta-analysis (*p=0.407)*. Additionally, use of the Duval and Tweedie’s trim-and-fill procedure revealed that removal of possible outlier studies did not substantially impact results of the meta-analysis (**Supplementary Figure 2)**. Taken together, these diagnostic procedures indicate that no single study was driving results, that publication biases were minimal, and that all ten studies should be included within the meta-analysis despite substantial between-study heterogeneity.

**Figure 2:**
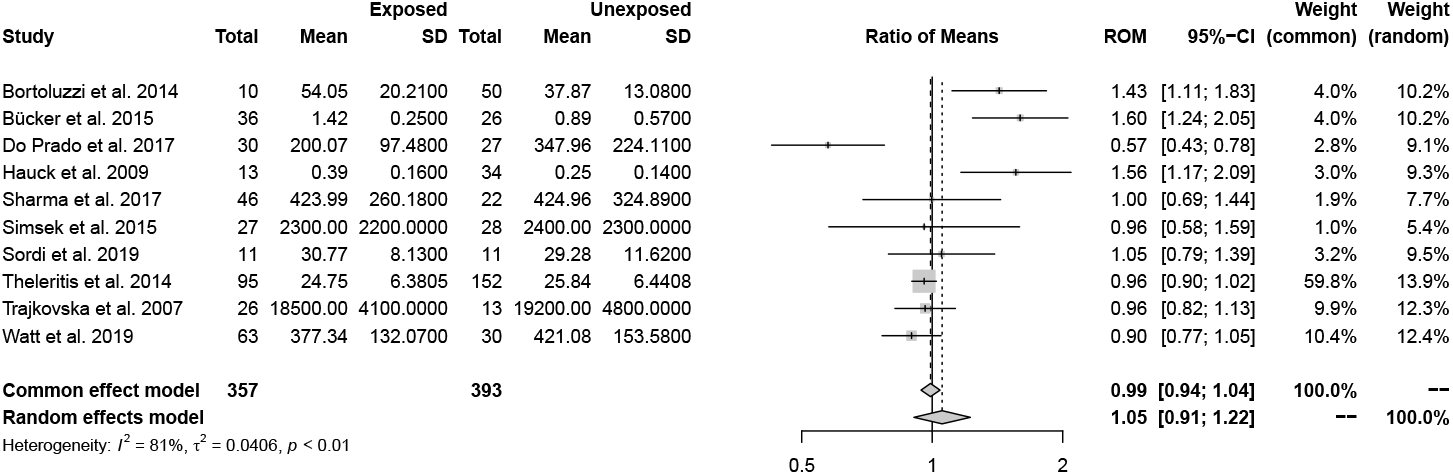
Forest plot evaluating ratio of mean BDNF protein levels between ACE-exposed and ACE-unexposed samples across all ten studies included within the meta-analysis. ROM: ratio of means 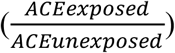. CI: Confidence Interval.

Studies were further analyzed by investigating the following sub-groups of interest: timing of BDNF measurement (childhood versus adulthood), childhood maltreatment as a specific subtype of ACE, and analyte in which BDNF was measured. To explore the impact of BDNF measurement timing, studies were grouped into two groups: BDNF measurement before the age of 20 and after the age of 20. Twenty years was identified as a cut-off based on 1) the specific demographics of samples from the included studies and 2) an interest in understanding the relationship of biomarker measurement during childhood and adolescence vs adulthood. In the subset of studies (*n=4*) measuring BDNF levels in subjects under the age of 20, BDNF levels were not significantly different in ACE-exposed versus ACE-unexposed individuals in either a fixed-effects (ROM: 1.15, 95% CI: 1.00, 1.33) or random-effects model (ROM: 1.07, 95% CI: 0.66, 1.74) (**Figure 3**). While significance was not noted, both models indicated higher BDNF levels within ACE-exposed subjects. Non-significant differences between the ACE-exposed and unexposed groups were also observed in the subset of studies (*n=6*) in which BDNF levels were measured after age 20, both in fixed-effects (ROM: 0.97, 95% CI: 0.92, 1.02) and random-effects models (ROM: 1.01, 95% CI: 0.90, 1.13) (**Figure 3**). No substantial heterogeneity in ROM estimates across the two age strata was observed in mixed-effects analysis (p=0.82).

**Figure 3:**
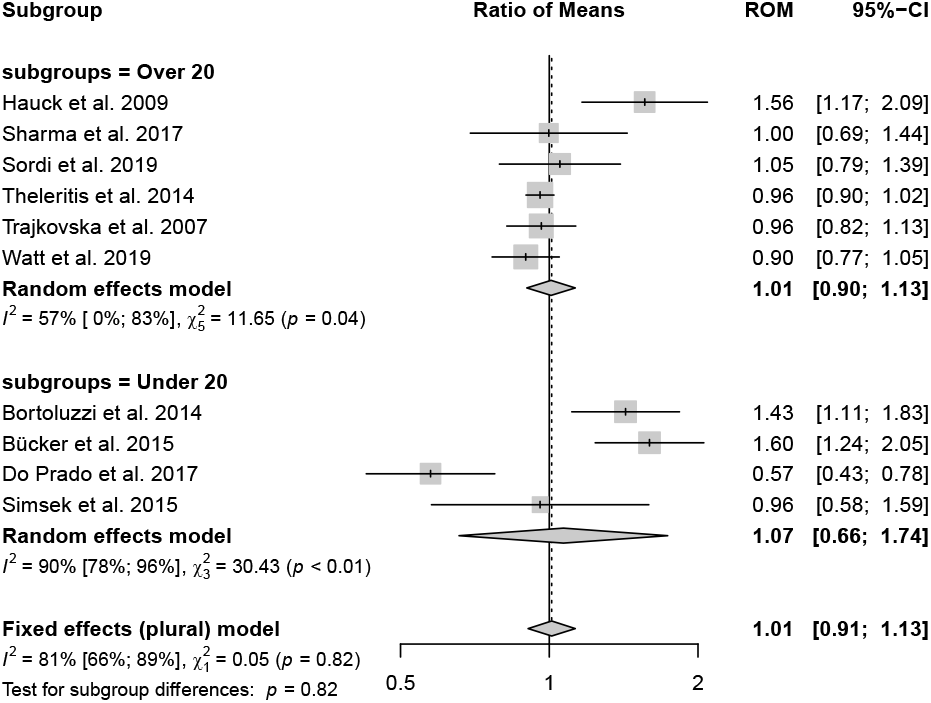
Forest plot evaluating ratio of mean BDNF protein levels between ACE-exposed and unexposed groups within subgroups investigating samples over and under the age of twenty. ROM: ratio of means 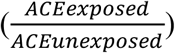. CI: Confidence Interval.

Limiting meta-analysis to the subgroup of studies specifically exploring childhood maltreatment as the ACE exposure (*n=5*), BDNF levels were non-significantly elevated in the ACE-exposed group in both fixed-effects (ROM: 1.13, 95% CI: 0.99,1.29) and random-effects models (ROM: 1.07; 95% CI: 0.74, 1.55) (**Figure 4**). Between-study heterogeneity was particularly high in this subset of studies (I^2^ = 87%). In addition to type of ACE and timing of measurements, the specific analyte assayed may also influence meta-analysis findings. Within the subset of studies quantifying serum levels of BDNF (*n=4*), BDNF levels were again non-significantly elevated in the ACE-exposed subjects in both fixed-effects (ROM: 1.09, 95% CI: 0.97, 1.22) and random-effects models (ROM 1.18, 95% CI: 0.88, 1.59) (**Figure 5**). Subset analysis of other analytes, such as plasma or blood, could not be performed due to the limited number of studies, and further investigation into tissue-specific alterations in BDNF signaling is needed to increase standardization and facilitate cross-study comparisons.

**Figure 4:**
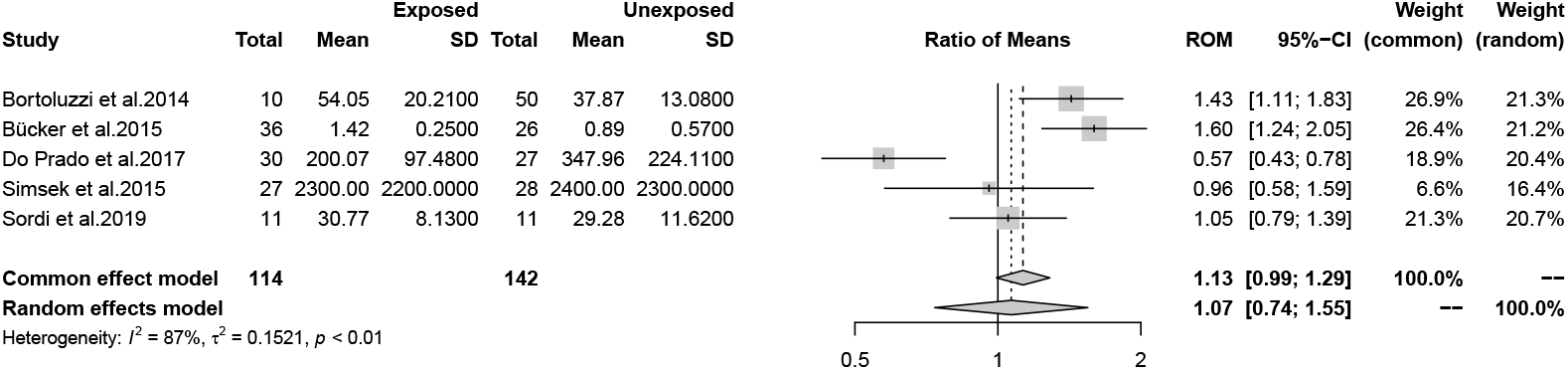
Forest plot evaluating ratio of mean BDNF protein levels between ACE-exposed and unexposed groups within studies investigating childhood maltreatment as the ACE exposure. ROM: ratio of means 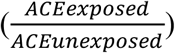. CI: Confidence Interval.

**Figure 5:**
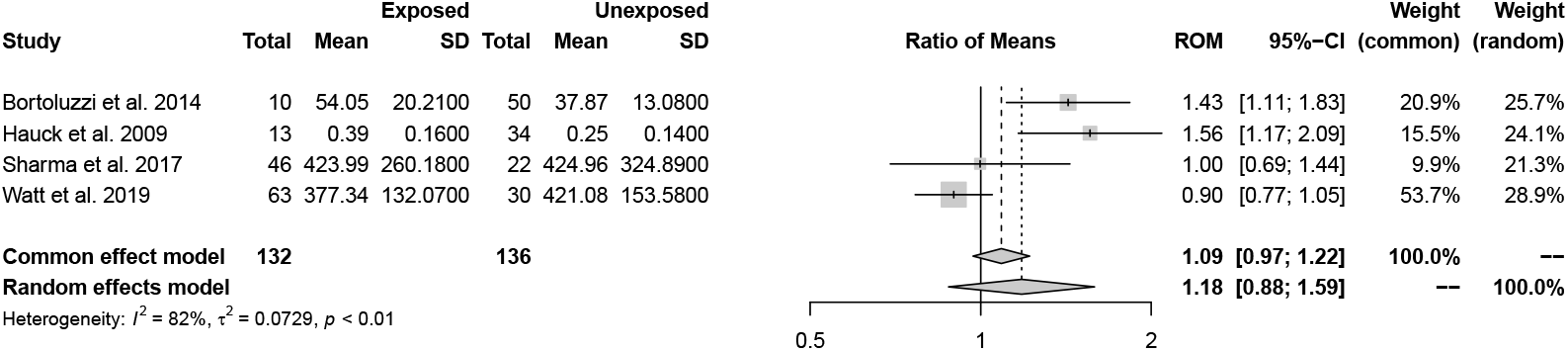
Forest plot of ratio evaluating mean BDNF protein levels between ACE-exposed and unexposed groups within studies measuring BDNF from the serum specifically. ROM: ratio of means 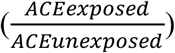. CI: Confidence Interval.

## Discussion

This is the first systematic review and meta-analysis exploring the impact of ACE exposure on protein levels of BDNF, a neural biomarker involved in childhood and adult neurogenesis and long-term memory formation. Results from the systematic review of twenty-two published studies indicate substantial heterogeneity in results, which may be related to differences in study design. Our systematic review revealed that studies in the field are largely restricted to clinical populations, primarily adults with various psychopathologies. The predominant study methodology remains measuring ACE exposure retrospectively during adulthood and assaying BDNF levels in these adult subjects, whereas a relative paucity of studies measured BDNF levels cross-sectionally during childhood and none have investigated this biomarker longitudinally. Studies also varied substantially in how ACE-exposed and ACE-unexposed participants were defined, which further contributes to a lack of consensus on the value of BDNF as a potential biomarker of ACE exposure. Findings from our meta-analysis of ten studies identified no statistically significant differences in BDNF protein levels between ACE-exposed and ACE-unexposed groups, although BDNF levels were generally elevated in ACE-exposed subjects in the subset of studies investigating childhood maltreatment as the specific type of ACE and this association approached significance in the fixed-effects model. Meta-analysis revealed substantial between-study heterogeneity, which likely contributed to the inconsistency of results across studies. Limiting analyses to specific age groups at the time of BDNF measurement or to specific analytes did not meaningfully resolve issues of between-study heterogeneity.

Within the ten studies undergoing formal synthesis through meta-analysis, clinical populations were predominant, being explored by six studies. However, all studies investigating clinical populations cannot be treated the same. Clinical populations encompassed by our meta-analysis included patients with a diagnosis of PTSD, first-episode psychosis, and major depressive disorder (MDD) (Bocchio-Chiavetto et al., 2010; Green, Corsi-Travali, & Neumeister, 2013; Pillai et al., 2010; Zhang et al., 2014). Both MDD and first-episode psychosis have been associated with lowered BDNF levels, while PTSD patients have been shown to display elevated levels of BDNF (Hsieh, Lin, Lee, & Huang, 2019; Mojtabavi, Saghazadeh, van den Heuvel, Bucker, & Rezaei, 2020; Pillai et al., 2010). Such relationships complicate the ability to draw firm conclusions about the effects of ACE exposure within subjects previously diagnosed with a psychopathology.

ACE exposure in our study was defined as exposure to *any* early-life stress exposure meeting 1998 CDC-Kaiser Study criteria, although others have alternatively defined it as having experienced more than three stressful life events (do Prado, Grassi-Oliveira, Daruy-Filho, Wieck, & Bauer, 2017; Trajkovska, Vinberg, Aznar, Knudsen, & Kessing, 2008). Similarly, definitions of the ACE-unexposed group in our study sample ranged from healthy controls to clinical patients with a psychiatric diagnosis who experienced fewer than three stressful life events. Utilization of varied ACE-exposure definitions may cloud results and is expected to limit precision of our meta-analytic point estimates. Future studies that adopt a more formally standardized ACE exposure definition would improve the field’s ability to better establish the relationship between ACE exposure and BDNF levels.

A previous meta-analysis that explored BDNF protein levels in patients with anxiety disorder, across a diversity of ACE exposures, also observed substantial between-study heterogeneity (Suliman, Hemmings, & Seedat, 2013). This may be partially attributable to the relatively few studies that specifically examined BDNF protein levels, the relatively small sample sizes within these studies, and the variation in study design that our own meta-analysis also encountered. Despite a strong rationale for the involvement of BDNF in mental health disorders, the possible link between ACE exposure and BDNF activity remains unclear, particularly in comparison to HPA biomarkers (*e.g*., cortisol) and inflammatory biomarkers (Kim et al., 2019).

Although outside the scope of our systematic review and meta-analysis of the effect of ACEs on BDNF protein levels, several studies have investigated ACE-associated changes in DNA methylation of the *BDNF* gene promoter region. Gene promoter methylation is a known epigenetic mechanism of regulating gene expression, and such an approach to understanding the relationship between ACEs and BDNF has both advantages and disadvantages compared with direct protein measurement, such as by ELISA. A major advantage to measuring ACE-associated changes in the dynamic methylation state of the *BDNF* gene promoter is that it includes a clear mechanism through which ACE exposure may modify BDNF activity. However, DNA methylation is strongly associated with differences in racial/ethnic ancestry and with age, so such studies need to explicitly account for such potential confounding factors in their analyses (Horvath, 2013; Kazmi et al., 2020) Furthermore, directly assaying BDNF protein levels would be expected to enhance cross-study comparisons and directly assesses what is likely a more physiologically-relevant endpoint. Indeed, while BDNF protein levels are perhaps more distal to the ACE exposure than methylation changes, they are more proximate to the clinical outcomes of greatest interest.

While our meta-analysis on all ten studies identified no significant association between ACE exposure and BDNF protein levels, subset analyses of more homogenous study designs revealed suggestive patterns. Subset analysis of studies exclusively investigating the impact of childhood maltreatment indicated a trend towards elevated BDNF levels in the ACE-exposed group, results which contrast with a prior study detailing decreases in BDNF protein levels and mRNA expression due to stressors and activation of the HPA axis (Bath, Schilit, & Lee, 2013). However, when looking at the specific studies in the subset analysis, heterogeneity persisted. Do Prado et al., the only study displaying a significant decrease in BDNF levels in the ACE-exposed group, measured only the impact of early-life stress. In contrast, the studies reporting significantly elevated levels of BDNF in ACE-exposed subjects had unique clinical samples with participants reporting foster care exposure and anxiety disorder onset. Thus, it may be that these additional factors modify the effects of ACE exposure on BDNF regulation in a different manner than is traditionally acknowledged within the field.

Two hypotheses have been investigated in relation to elevated BDNF levels in the face of stress exposure. One hypothesis attributes increased BDNF protein levels to compensatory effect, wherein stress exposure leads to increased BDNF levels in order to accommodate for stress-related decreases in neurogenesis (Greisen, Altar, Bolwig, Whitehead, & Wörtwein, 2005). A second hypothesis considers the form of BDNF that is being upregulated or downregulated. BDNF can be present as proBDNF or mature-BDNF, and these forms have opposing functions. Pro-BDNF has been found to induce apoptosis via interaction with the inflammatory system, while mature-BDNF contributes to synaptogenesis and memory consolidation (Mojtabavi et al., 2020). An increase in pro-BDNF levels specifically could be detrimental due to its contribution to neuronal death. Commercially available ELISA assays for BDNF typically measure mature-BDNF unless explicitly specified to measure pro-BDNF, however, it remains possible that a subset of studies included in our meta-analysis measured pro-BDNF without clearly stating so.

Subgroup analysis of BDNF levels in groups under and over the age of 20 also indicates possible time dependency in changes of BDNF protein levels. While there was no significant difference between the protein levels of BDNF between the two groups in our mixed-effects model, there was a stronger trend towards higher BDNF protein levels measured in the ACE-exposed group prior to the age of 20. The highest levels of BDNF mRNA and protein levels are found during the adolescent/young adult period, after which levels are found to decrease during middle to late adulthood (Katoh-Semba et al., 2007). Our results may reflect developmental changes in BDNF protein levels, or a waning effect of ACE-exposure on BDNF at more removed timepoints. BDNF levels may also be elevated following ACE exposure due to the previously mentioned compensatory mechanism, but such changes may not persist longitudinally. Exposure to a chronic stress-state has been found to be associated with lowered BDNF levels in murine models (Algamal et al., 2018). ACE exposure can result in persistent experiences of chronic stressors due to the longitudinal impacts of ACEs, as children exposed to parental substance abuse or sexual abuse have been found to engage in riskier future behaviors such as engaging in abusive relationships or pursuing risky sexual behaviors, but teasing out these effects across diverse studies designs with varied ACE exposures remains difficult (Lalor & McElvaney, 2010; Senn, Carey, & Vanable, 2008).

If ACE exposure increases the likelihood of chronic stress exposure and these jointly lead to BDNF blunting in adulthood, such a mechanism has particular relevance given the increased susceptibility of children with ACE exposure to the development of mental disorders during adulthood. Decreased BDNF levels have been associated with MDD, anxiety, and schizophrenia, all diagnoses seen at higher rates in ACE exposed adults (Hsieh et al., 2019). Changes in BDNF levels could thus link ACE exposure to poorer mental health outcomes. Future studies should focus on both further investigating developmental changes in BDNF levels and also on measuring lifelong stress in addition to ACE exposure in order to evaluate the impact of chronic stress on BDNF blunting.

Several limitations must be acknowledged within this study. First, several publications did not report the true mean BDNF values within their samples, instead displaying the data graphically or reporting relevant statistical analysis results. Authors were contacted, and studies were excluded if data were not provided. Inclusion of these studies would have been beneficial given the small sample size of the overall meta-analysis and the sub-group meta-analyses. It would be beneficial for future studies to provide raw biomarker data in order to facilitate cross-study comparisons and meta-analyses. Provision of all relevant data would be in-line with the general shift towards data transparency in health research and the increased willingness of journals to permit the inclusion of online-only supplemental data files and tables. Second, the small sample size (<10) of the subset analyses prevented evaluation of publication bias via the Egger’s test, and results may also be unduly influenced by a single study. Additionally, the small sample size of studies included within the meta-analysis prevented analysis of all possible sub-groups, such as exploring the impact of ACEs other than childhood maltreatment or differential impact of other analytes (*e.g*., whole blood or plasma) on BDNF protein levels. Relationships between these variables and BDNF should not be disregarded and instead be further investigated in order to determine mechanistic details of putative ACE-mediated dysregulation of BDNF. Increased data reporting along with further investigation into the ACE-BDNF relationship would attenuate these limitations and help to better elucidate the impact of ACEs on BDNF protein levels specifically. Future investigations could also consider expanding ACE exposures analyzed in relation to BDNF levels, moving beyond exploring the impact of traditional ACEs, such as parental mental health or parental substance abuse, and towards acknowledging an expanded list of ACE exposures, such as experiences of discrimination or exposure to violent environments (Afifi, 2020; Cronholm et al., 2015). Diversification of the ACE exposures considered in addition to explicit sub-analysis of the traditional ACEs and BDNF protein levels will ensure a more thorough understanding of the ACE-BDNF relationship that reflects unique stressors present in the environment today.

In summary, while individual studies provide evidence of ACE-dependent changes in BDNF protein levels, limited sample sizes and heterogeneous study designs limit the ability to reach a consensus on the strength and breadth of this possible relationship. Differential regulation of BDNF levels based on the type of ACE exposure, the acute versus chronic nature of the ACE exposure, and the timing of biomarker measurement seem likely. Our meta-analysis was purposefully broad in nature to begin the process of consolidating the state of the field and to investigate the overarching relationship of ACE exposure and BDNF protein levels. While it was limited by heterogeneity and the small sample sizes used by individual studies, it can serve as a starting point to inform the design of future studies. Understanding the complex ACE-BDNF relationship will enable greater insight into how ACE exposure may predispose ACE-exposed children towards poorer mental health outcomes, and thus may help nominate treatment approaches to mitigate mental health risks in ACE-exposed children.

## Data Availability

All data produced in the present study are available upon request to the authors.

## Acknowledgements

We thank Jillian Hurst and the Duke Children’s Health and Discovery Initiative for their support through this project. We thank Rick Hoyle for his review of the manuscript, and detailed suggestions for improvement. We are also grateful to Sallie Permar for launching the Children’s Health and Discovery Initiative scholars program and inviting undergraduates to participate in pediatric research projects.

## Works Cited

Afifi, T. O. (2020). Considerations for expanding the definition of ACEs. In Adverse childhood experiences (pp. 35–44): Elsevier.

Algamal, M., Ojo, J. O., Lungmus, C. P., Muza, P., Cammarata, C., Owens, M. J., … Crawford, F. (2018). Chronic hippocampal abnormalities and blunted HPA axis in an animal model of repeated unpredictable stress. Frontiers in behavioral neuroscience, 12, 150.

Bath, K., Schilit, A., & Lee, F. (2013). Stress effects on BDNF expression: effects of age, sex, and form of stress. Neuroscience, 239, 149–156.

Baumeister, D., Akhtar, R., Ciufolini, S., Pariante, C. M., & Mondelli, V. (2016). Childhood trauma and adulthood inflammation: a meta-analysis of peripheral C-reactive protein, interleukin-6 and tumour necrosis factor-α. Mol Psychiatry, 21(5), 642–649. doi:10.1038/mp.2015.67

Bekinschtein, P., Cammarota, M., Izquierdo, I., & Medina, J. H. (2007). Reviews: BDNF and Memory Formation and Storage. The Neuroscientist, 14(2), 147–156. doi:10.1177/1073858407305850

Benedetti, F., Ambree, O., Locatelli, C., Lorenzi, C., Poletti, S., Colombo, C., & Arolt, V. (2017). The effect of childhood trauma on serum BDNF in bipolar depression is modulated by the serotonin promoter genotype. Neurosci Lett, 656, 177–181.

Binder, D. K., & Scharfman, H. E. (2004). Brain-derived neurotrophic factor. Growth factors (Chur, Switzerland), 22(3), 123–131. doi:10.1080/08977190410001723308

Bocchio-Chiavetto, L., Bagnardi, V., Zanardini, R., Molteni, R., Gabriela Nielsen, M., Placentino, A., … Riva, M. A. (2010). Serum and plasma BDNF levels in major depression: a replication study and meta-analyses. The World Journal of Biological Psychiatry, 11(6), 763–773.

Bortoluzzi, A., Salum, G. A., Blaya, C., Silveira, P. P., Grassi-Oliveira, R., da Rosa, E. D., … Schuch, I. (2014). Mineralocorticoid receptor genotype moderates the association between physical neglect and serum BDNF. Journal of psychiatric research, 59, 8–13.

Bücker, J., Fries, G., Kapczinski, F., Post, R., Yatham, L., Vianna, P., … Aguiar, B. (2015). Brain-derived neurotrophic factor and inflammatory markers in school-aged children with early trauma. Acta Psychiatrica Scandinavica, 131(5), 360–368.

Buss, C., Lord, C., Wadiwalla, M., Hellhammer, D. H., Lupien, S. J., Meaney, M. J., & Pruessner, J. C. (2007). Maternal care modulates the relationship between prenatal risk and hippocampal volume in women but not in men. Journal of Neuroscience, 27(10), 2592–2595.

Calabrese, F., Rossetti, A. C., Racagni, G., Gass, P., Riva, M. A., & Molteni, R. (2014). Brain-derived neurotrophic factor: a bridge between inflammation and neuroplasticity. Frontiers in Cellular Neuroscience, 8(430). doi:10.3389/fncel.2014.00430

Cronholm, P. F., Forke, C. M., Wade, R., Bair-Merritt, M. H., Davis, M., Harkins-Schwarz, M., … Fein, J. A. (2015). Adverse childhood experiences: Expanding the concept of adversity. American journal of preventive medicine, 49(3), 354–361.

Deighton, S., Neville, A., Pusch, D., & Dobson, K. (2018). Biomarkers of adverse childhood experiences: A scoping review. Psychiatry Res, 269, 719–732.

Dempster, K. S., O’Leary, D. D., MacNeil, A. J., Hodges, G. J., & Wade, T. J. (2020). Linking the hemodynamic consequences of adverse childhood experiences to an altered HPA axis and acute stress response. Brain, behavior, and immunity.

Dempster, K. S., O’Leary, D. D., MacNeil, A. J., Hodges, G. J., & Wade, T. J. (2021). Linking the hemodynamic consequences of adverse childhood experiences to an altered HPA axis and acute stress response. Brain Behav Immun, 93, 254–263. doi:10.1016/j.bbi.2020.12.018

do Prado, C. H., Grassi-Oliveira, R., Daruy-Filho, L., Wieck, A., & Bauer, M. E. (2017). Evidence for immune activation and resistance to glucocorticoids following childhood maltreatment in adolescents without psychopathology. Neuropsychopharmacology, 42(11), 2272–2282.

Felitti, V. J., Anda, R. F., Nordenberg, D., Williamson, D. F., Spitz, A. M., Edwards, V., … Marks, J. S. (1998). Relationship of childhood abuse and household dysfunction to many of the leading causes of death in adults. The Adverse Childhood Experiences (ACE) Study. Am J Prev Med, 14(4), 245–258. doi:10.1016/s0749-3797(98)00017-8

Gaglia, R. J. (2021). Brain-derived neurotrophic factor (BDNF), B-catenin, and cortisol levels correlated with the severity of adverse childhood experiences (ACEs) score in patients with Schizophrenia spectrum disorders.

Godoy, L. C., Frankfurter, C., Cooper, M., Lay, C., Maunder, R., & Farkouh, M. E. (2021). Association of adverse childhood experiences with cardiovascular disease later in life: a review. JAMA cardiology, 6(2), 228–235.

Green, C. R., Corsi-Travali, S., & Neumeister, A. (2013). The role of BDNF-TrkB signaling in the pathogenesis of PTSD. Journal of depression & anxiety, 2013(S4).

Greisen, M. H., Altar, C. A., Bolwig, T. G., Whitehead, R., & Wörtwein, G. (2005). Increased adult hippocampal brain-derived neurotrophic factor and normal levels of neurogenesis in maternal separation rats. J Neurosci Res, 79(6), 772–778.

Grummitt, L. R., Kreski, N. T., Kim, S. G., Platt, J., Keyes, K. M., & McLaughlin, K. A. (2021). Association of Childhood Adversity With Morbidity and Mortality in US Adults: A Systematic Review. JAMA Pediatr, 175(12), 1269–1278. doi:10.1001/jamapediatrics.2021.2320

Hauck, S., Kapczinski, F., Roesler, R., de Moura Silveira Jr, É., Magalhães, P. V., Kruel, L. R. P., … Ceitlin, L. H. F. (2010). Serum brain-derived neurotrophic factor in patients with trauma psychopathology. Progress in Neuro-Psychopharmacology and Biological Psychiatry, 34(3), 459–462.

Horvath, S. (2013). DNA methylation age of human tissues and cell types. Genome biology, 14(10), 1–20.

Hsieh, M.-T., Lin, C.-C., Lee, C.-T., & Huang, T.-L. (2019). Abnormal brain-derived neurotrophic factor exon IX promoter methylation, protein, and mRNA levels in patients with major depressive disorder. Journal of Clinical Medicine, 8(5), 568.

Hughes, K., Bellis, M. A., Hardcastle, K. A., Sethi, D., Butchart, A., Mikton, C., … Dunne, M. P. (2017). The effect of multiple adverse childhood experiences on health: a systematic review and meta-analysis. The Lancet Public Health, 2(8), e356–e366.

Jiang, S., Postovit, L., Cattaneo, A., Binder, E. B., & Aitchison, K. J. (2019). Epigenetic modifications in stress response genes associated with childhood trauma. Frontiers in psychiatry, 10, 808.

Jin, Y., Sun, L. H., Yang, W., Cui, R. J., & Xu, S. B. (2019). The role of BDNF in the neuroimmune axis regulation of mood disorders. Frontiers in neurology, 10, 515.

Kalmakis, K. A., & Chandler, G. E. (2015). Health consequences of adverse childhood experiences: A systematic review. Journal of the American Association of Nurse Practitioners, 27(8), 457–465.

Katoh-Semba, R., Wakako, R., Komori, T., Shigemi, H., Miyazaki, N., Ito, H., … Yoshida, F. (2007). Age-related changes in BDNF protein levels in human serum: differences between autism cases and normal controls. International Journal of Developmental Neuroscience, 25(6), 367–372.

Kauer-Sant’Anna, M., Tramontina, J., Andreazza, A. C., Cereser, K., Costa, S. d., Santin, A., … Kapczinski, F. (2007). Traumatic life events in bipolar disorder: impact on BDNF levels and psychopathology. Bipolar Disorders, 9, 128–135.

Kazmi, N., Elliott, H. R., Burrows, K., Tillin, T., Hughes, A. D., Chaturvedi, N., … Relton, C. L. (2020). Associations between high blood pressure and DNA methylation. PLoS One, 15(1), e0227728.

Kim, S., Watt, T., Ceballos, N., & Sharma, S. (2019). Adverse childhood experiences and neuroinflammatory biomarkers—The role of sex. Stress and Health, 35(4), 432–440.

Kraneveld, A. D., de Theije, C. G., van Heesch, F., Borre, Y., de Kivit, S., Olivier, B., … Garssen, J. (2014). The neuro-immune axis: Prospect for novel treatments for mental disorders. Basic & clinical pharmacology & toxicology, 114(1), 128–136.

Kuhlman, K. R., Horn, S. R., Chiang, J. J., & Bower, J. E. (2020). Early life adversity exposure and circulating markers of inflammation in children and adolescents: A systematic review and meta-analysis. Brain Behav Immun, 86, 30–42. doi:10.1016/j.bbi.2019.04.028

Lalor, K., & McElvaney, R. (2010). Child sexual abuse, links to later sexual exploitation/high-risk sexual behavior, and prevention/treatment programs. Trauma, Violence, & Abuse, 11(4), 159–177.

Merrick, M. T., Ford, D. C., Ports, K. A., Guinn, A. S., Chen, J., Klevens, J., … Daniel, V. M. (2019). Vital signs: estimated proportion of adult health problems attributable to adverse childhood experiences and implications for prevention—25 states, 2015–2017. Morbidity and Mortality Weekly Report, 68(44), 999.

Mojtabavi, H., Saghazadeh, A., van den Heuvel, L., Bucker, J., & Rezaei, N. (2020). Peripheral blood levels of brain-derived neurotrophic factor in patients with post-traumatic stress disorder (PTSD): A systematic review and meta-analysis. PLoS One, 15(11), e0241928.

Naert, G., Ixart, G., Maurice, T., Tapia-Arancibia, L., & Givalois, L. (2011). Brain-derived neurotrophic factor and hypothalamic-pituitary-adrenal axis adaptation processes in a depressive-like state induced by chronic restraint stress. Mol Cell Neurosci, 46(1), 55–66. doi:10.1016/j.mcn.2010.08.006

Navalta, C. P., McGee, L., & Underwood, J. (2018). Adverse childhood experiences, brain development, and mental health: A call for neurocounseling. Journal of Mental Health Counseling, 40(3), 266–278.

Neves, I., Dinis-Oliveira, R. J., & Magalhães, T. (2019). Epigenomic mediation after adverse childhood experiences: a systematic review and meta-analysis. Forensic Sciences Research, 1–12.

Petruccelli, K., Davis, J., & Berman, T. (2019). Adverse childhood experiences and associated health outcomes: A systematic review and meta-analysis. Child abuse & neglect, 97, 104127.

Pilkay, S. R., Combs-Orme, T., Tylavsky, F., Bush, N., & Smith, A. K. (2020). Maternal trauma and fear history predict BDNF methylation and gene expression in newborns. PeerJ, 8, e8858.

Pillai, A., Kale, A., Joshi, S., Naphade, N., Raju, M., Nasrallah, H., & Mahadik, S. P. (2010). Decreased BDNF levels in CSF of drug-naive first-episode psychotic subjects: correlation with plasma BDNF and psychopathology. International Journal of Neuropsychopharmacology, 13(4), 535–539.

Pretty, C., D O’Leary, D., Cairney, J., & Wade, T. J. (2013). Adverse childhood experiences and the cardiovascular health of children: a cross-sectional study. BMC pediatrics, 13(1), 1–8.

Schauss, E., Horn, G., Ellmo, F., Reeves, T., Zettler, H., Bartelli, D., … West, S. (2019). Fostering intrinsic resilience: A neuroscience-informed model of conceptualizing and treating adverse childhood experiences. Journal of Mental Health Counseling, 41(3), 242–259.

Senn, T. E., Carey, M. P., & Vanable, P. A. (2008). Childhood and adolescent sexual abuse and subsequent sexual risk behavior: Evidence from controlled studies, methodological critique, and suggestions for research. Clinical psychology review, 28(5), 711–735.

Sharma, S., Graham, R., Rohde, R., & Ceballos, N. A. (2017). Stress-induced change in serum BDNF is related to quantitative family history of alcohol use disorder and age at first alcohol use. Pharmacology Biochemistry and Behavior, 153, 12–17.

Simsek, S., Uysal, C., Kaplan, I., Yuksel, T., & Aktas, H. (2015). BDNF and cortisol levels in children with or without post-traumatic stress disorder after sustaining sexual abuse. Psychoneuroendocrinology, 56, 45–51.

Sordi, A. O., von Diemen, L., Kessler, F. H., Schuch, S., Ornell, F., Kapczinski, F., … Salum, G. A. (2019). Effects of childhood trauma on BDNF and TBARS during crack-cocaine withdrawal. Brazilian Journal of Psychiatry, 42, 214–217.

Suliman, S., Hemmings, S. M., & Seedat, S. (2013). Brain-Derived Neurotrophic Factor (BDNF) protein levels in anxiety disorders: systematic review and meta-regression analysis. Frontiers in integrative neuroscience, 7, 55.

Theleritis, C., Fisher, H. L., Shäfer, I., Winters, L., Stahl, D., Morgan, C., … Vitoratou, S. (2014). Brain derived neurotropic factor (BDNF) is associated with childhood abuse but not cognitive domains in first episode psychosis. Schizophr Res, 159(1), 56–61.

Trajkovska, V., Vinberg, M., Aznar, S., Knudsen, G. M., & Kessing, L. V. (2008). Whole blood BDNF levels in healthy twins discordant for affective disorder: association to life events and neuroticism. Journal of affective disorders, 108(1-2), 165–169.

Tsai, S.-J. (2018). Critical issues in BDNF Val66Met genetic studies of neuropsychiatric disorders. Frontiers in molecular neuroscience, 11, 156.

Watt, T., Ceballos, N., Kim, S., Pan, X., & Sharma, S. (2020). The unique nature of depression and anxiety among college students with adverse childhood experiences. Journal of child & adolescent trauma, 13(2), 163–172.

Wilson, M., & Perez Vallejos, E. (2021). The Role of Neuroscience in the Effects of Adverse Childhood Experiences in Relation to Risk Taking, with Specific Reference to Risk Assessment During a Pandemic, a Review of the Literature. Available at SSRN 3865103.

Zhang, L., Benedek, D., Fullerton, C., Forsten, R., Naifeh, J., Li, X., … Xing, G. (2014). PTSD risk is associated with BDNF Val66Met and BDNF overexpression. Molecular psychiatry, 19(1), 8–10.

