## Supplementary figures and images for "Systematic Review and Meta-Analysis of the Effect of Adverse Childhood Experiences (ACEs) on Brain-Derived Neurotrophic Factor (BDNF) Levels"

### Supplementary Figure 1

**Supplementary Figure 1: Sample REDCap Data Collection Tool**

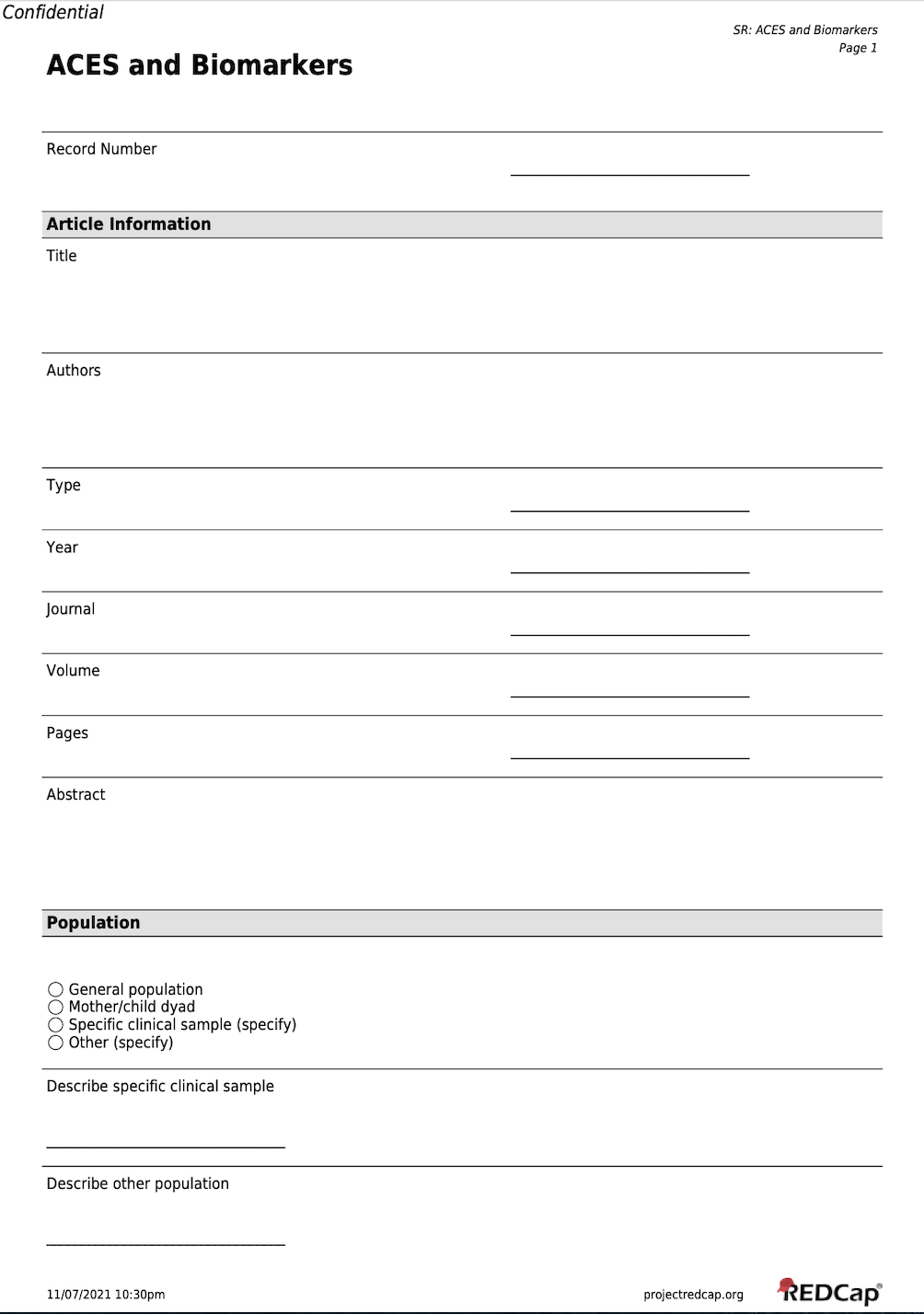

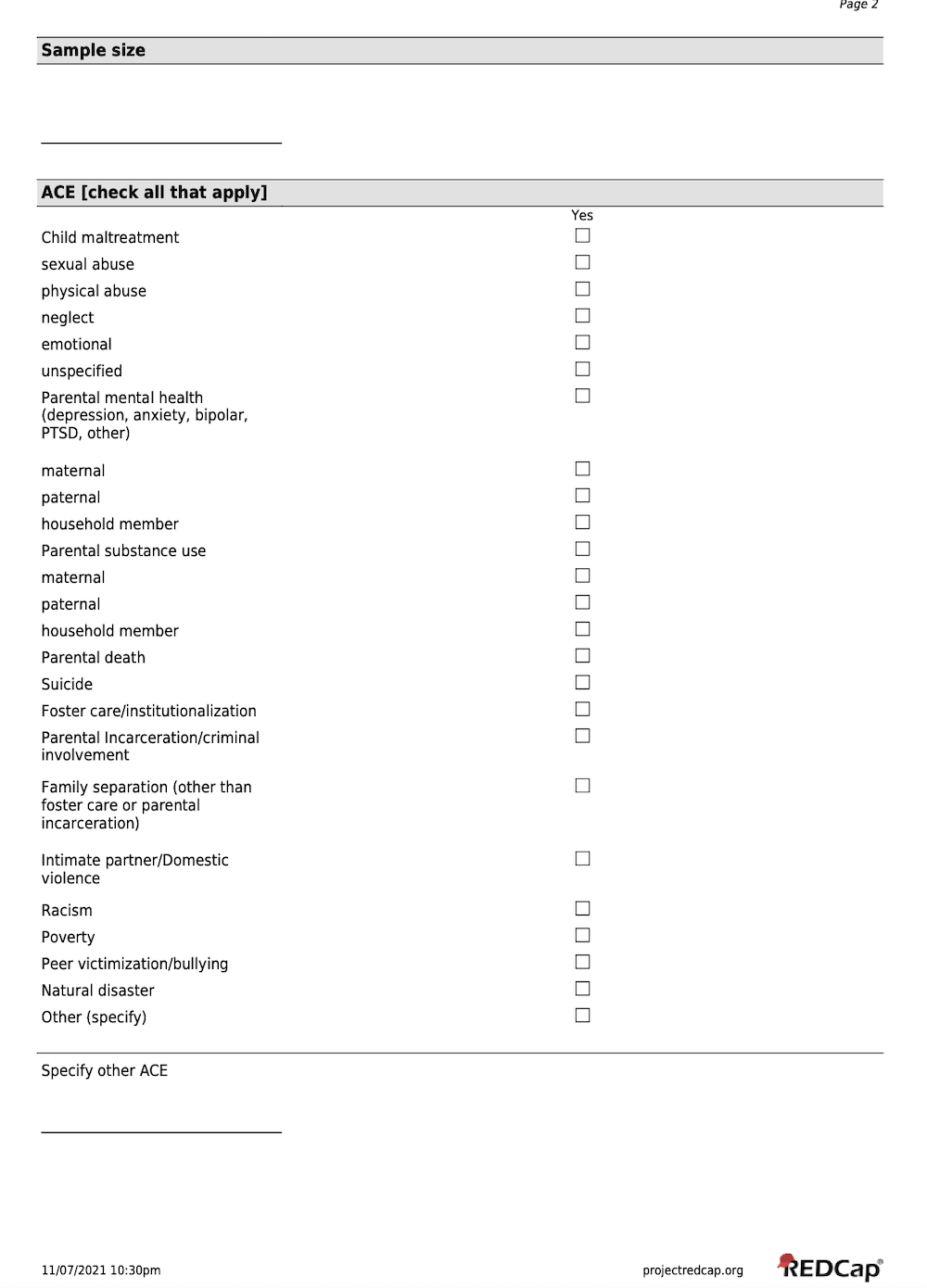

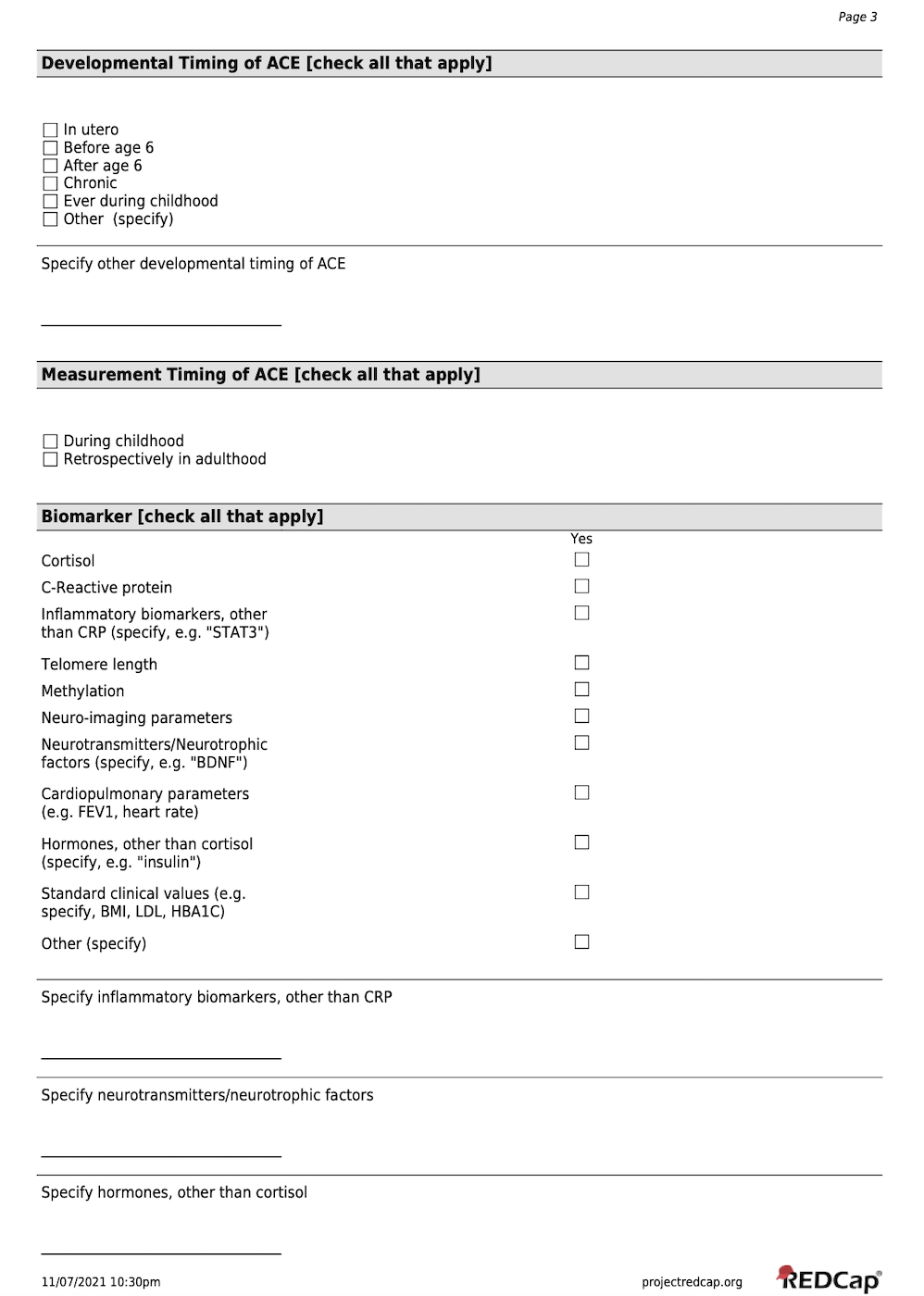

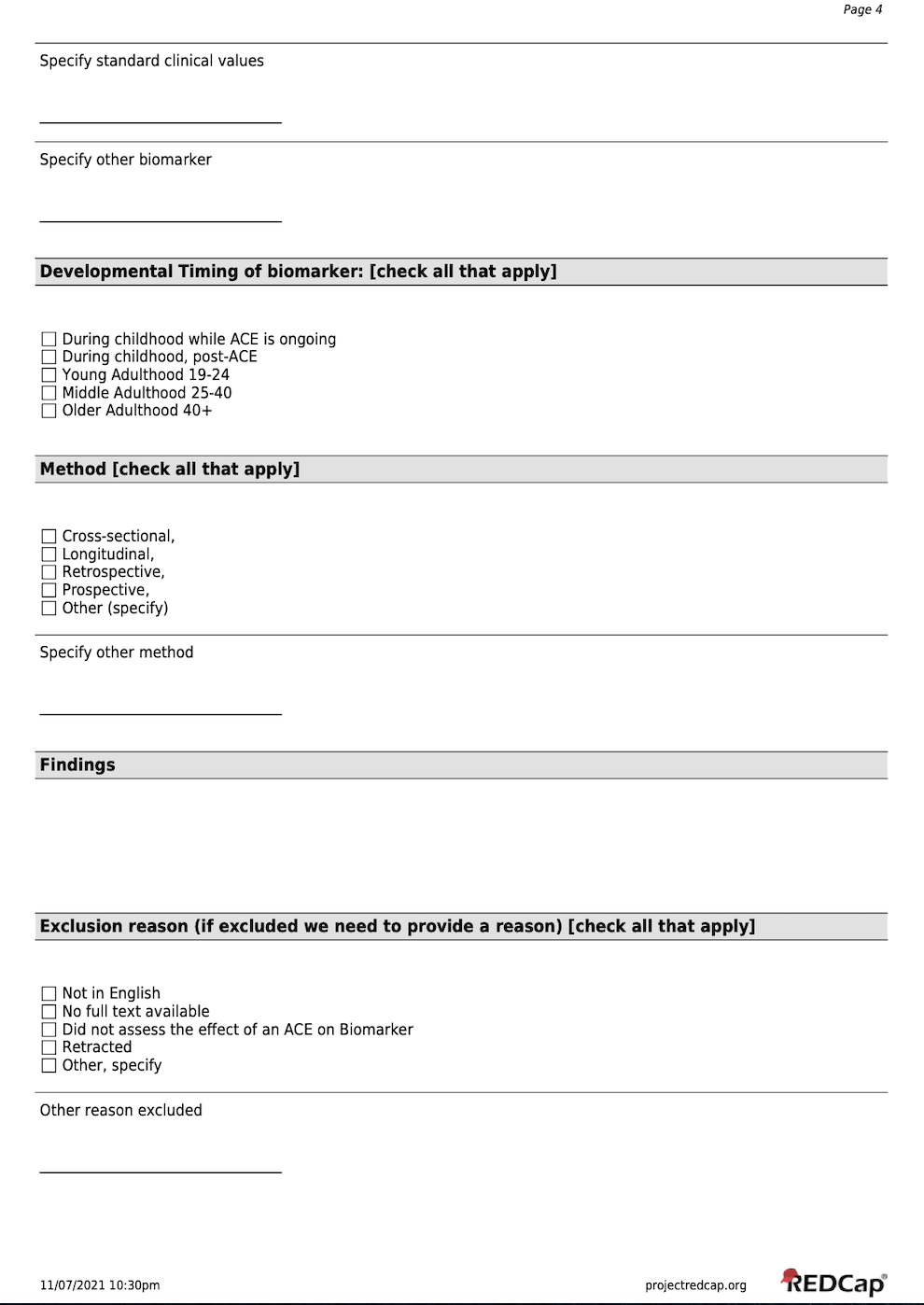

### Supplementary Figure 2

**Supplementary Figure 2: Duval and Tweedie’s Funnel Plot**

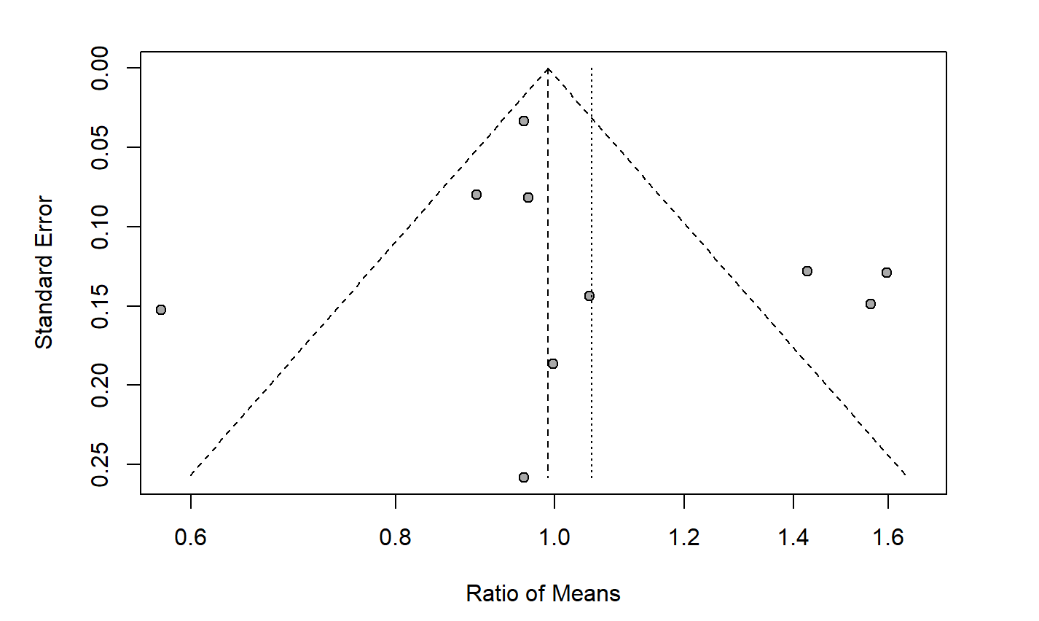
