## Supplementary Table 1 for "Systematic Review and Meta-Analysis of the Effect of Adverse Childhood Experiences (ACEs) on Brain-Derived Neurotrophic Factor (BDNF) Levels"

**Supplementary Table 1: Search Strategy Report**

Topic: How does experiencing adverse childhood experience changes children’s biologic reaction to stress and other biomarkers of health.

Searcher: SJK

Date: 4.27.2020

Database (including vendor/platform): Ovid/Medline

| Net | Set |  | Results |
| --- | --- | --- | --- |
| ACEs | 1 | exp "Adult Survivors of Child Adverse Events"/ or exp Adverse Childhood Experiences/ | 3333 |
|  | 2 | exp Child Welfare/ OR exp "Child of Impaired Parents"/ OR exp Child Abuse/ or exp "Adult Survivors of Child Abuse"/ OR exp Child Abuse, Sexual/ OR exp Physical Abuse/ OR exp Substance-Related Disorders/ OR exp Domestic Violence/ or exp Divorce/ OR exp Family Separation/ OR exp Poverty/ OR exp Racism/ or exp Suicide/ or exp Intimate Partner Violence/ or exp Depressive Disorder | 540707 |
|  | 3 | (ACEs or (toxic adj1 stress) or (childhood adj1 adversity) or (childhood trauma) or (adverse adj1 childhood) or (childhood adj1 adversities) or ((Adverse or adversity or adversities) adj3 (child or childhood or children) adj3 (exposure or exposures or exposed or event or events or experience or experiences or experienced or experiencing))).ti,ab. | 7888 |
|  | 4 | 1 OR 2 OR 3 | 545608 |
| Biomarkers | 5 | exp Biomarkers/ | 735410 |
|  | 6 | (Biomarker or biomarkers or epigenomics OR epigenetic OR epigenetics OR interleukins OR interleukin OR TNF OR interferon OR interferons OR fibrinogen OR leukocyte OR leukocytes OR cytokine OR cytokines OR chemokines OR chemokine OR insulin OR telomere OR cortisol OR telomeres OR (Tumor adj1 Necrosis adj1 Factor adj1 alpha) OR (c adj1 reactive adj1 protein) OR CRP OR telomeres OR telomere or ((Biological OR bio OR immune OR serum OR plasma OR blood OR saliva OR urine OR laboratory OR viral OR clinical OR surrogate OR immunologic OR biochemical or molecular or genetic or inflammatory OR inflammation OR physiological OR cardiometabolic OR cardiovascular OR proinflammatory) adj2 (Marker OR markers OR endpoint OR levels OR level))).ti,ab | 2085320 |
|  | 7 | 5 or 6 | 2537119 |
|  | 8 | 4 and 7 | 26947 |
| Stress reaction | 9 | exp Stress, Psychological/ or exp Stress, Physiological/ OR exp Stress Disorders, Post-Traumatic/ or exp Stress Disorders, Traumatic/ or exp Adaptation, Biological/ or exp Adaptation, Psychological/ or exp Adaptation, Physiological/ or exp Cognitive Dysfunction/ or exp Life Change Events/ OR exp Allostasis/ | 627568 |
|  | 10 | (reaction or reactions or reacted or reactive or reacting or stress or stresses or stressor or stressors or stressed or adaptation or adaption or adapt or adapted or adapts or adapting or development or developing or developed or develop or develops or developmental or maladaptation or maladaption or maladapted or outcome or outcomes or impact or impacts or influence or influences or function or functions or functioning or functioned OR allostasis OR (allostatic adj1 load)).ti,ab | 10147768 |
|  | 11 | 9 or 10 | 10336323 |
|  | 12 | 8 and 11 | 14520 |
| Children | 13 | exp child/ or exp child, preschool/ or exp infant/ or exp Adolescent/ | 3524617 |
|  | 14 | (child or children or childhood or pediatric or pediatrics or paediatric or paediatrics or kid or kids or boy or girl or boys or girls or infant or infants or infancy or neonate or neonates or baby or babies or juvenile or juveniles or adolescent or adolescents or adolescence or teen or teens or teenager or teenagers or boyhood or girlhood or newborn or newborns).ti,ab | 2189417 |
|  | 15 | 13 or 14 | 4132033 |
|  | 16 | 12 and 15 | 3940 |
|  | 17 | 16 not (case reports or editorial or letter or comment).pt. | 3809 |
|  | 18 | 17 not (Animals/ not (Animals/ and Humans/)) | 3686 |
|  |  | (25783196 OR 30605796 OR 27647050 OR 27275737 OR 25496803 OR 30067291).ui | Catches 3 |

Topic: How does experiencing adverse childhood experience changes children’s biologic reaction to stress and other biomarkers of health.

Searcher: SJK

Date: 4.28.2020

Database (including vendor/platform): Embase

| Set |  | Results |
| --- | --- | --- |
| 1 | 'child welfare'/exp OR 'child of impaired parents'/exp OR 'child abuse'/exp OR 'childhood trauma survivor'/exp OR 'child sexual abuse'/exp OR 'physical abuse'/exp OR 'drug dependence'/exp OR 'domestic violence'/exp OR 'divorce'/exp OR 'family separation'/exp OR 'poverty'/exp OR 'racism'/exp OR 'suicide'/exp OR 'partner violence'/exp OR 'depression'/exp | 888108 |
| 2 | (ACEs OR (toxic NEAR/1 stress) OR (childhood NEAR/1 adversity) OR ("childhood trauma") OR (adverse NEAR/1 childhood) OR (childhood NEAR/1 adversities) OR ((Adverse OR adversity OR adversities) NEAR/3 (child OR childhood OR children) NEAR/3 (exposure OR exposures OR exposed OR event OR events OR experience OR experiences OR experienced OR experiencing))):ti,ab | 11124 |
| 3 | 1 OR 2 | 893335 |
| 4 | 'biological marker'/exp | 298625 |
| 5 | (Biomarker OR biomarkers OR epigenomics OR epigenetic OR epigenetics OR interleukins OR interleukin OR TNF OR interferon OR interferons OR fibrinogen OR leukocyte OR leukocytes OR cytokine OR cytokines OR chemokines OR chemokine OR insulin OR telomere OR cortisol OR telomeres OR (Tumor NEAR/1 Necrosis NEAR/1 Factor NEAR/1 alpha) OR (c NEAR/1 reactive NEAR/1 protein) OR CRP OR telomeres OR telomere OR ((Biological OR bio OR immune OR serum OR plasma OR blood OR saliva OR urine OR laboratory OR viral OR clinical OR surrogate OR immunologic OR biochemical OR molecular OR genetic OR inflammatory OR inflammation OR physiological OR cardiometabolic OR cardiovascular OR proinflammatory) NEAR/2 (Marker OR markers OR endpoint OR levels OR level))):ti,ab | 2907043 |
| 6 | 4 or 5 | 2970217 |
| 7 | 3 and 6 | 57001 |
| 8 | 'mental stress'/exp OR 'physiological stress'/exp OR 'posttraumatic stress disorder'/exp OR 'adaptation'/exp OR 'cognitive defect'/exp OR 'life event'/exp OR 'allostasis'/exp | 835833 |
| 9 | (reaction OR reactions OR reacted OR reactive OR reacting OR stress OR stresses OR stressor OR stressors OR stressed OR adaptation OR adaption OR adapt OR adapted OR adapts OR adapting OR development OR developing OR developed OR develop OR develops OR developmental OR maladaptation OR maladaption OR maladapted OR outcome OR outcomes OR impact OR impacts OR influence OR influences OR function OR functions OR functioning OR functioned OR allostasis OR (allostatic NEAR/1 load)):ti,ab | 13451136 |
| 10 | 8 or 9 | 13774791 |
| 11 | 10 and 7 | 36799 |
| 12 | 'pediatrics'/exp OR 'infant'/exp OR 'child'/exp OR 'adolescent'/exp OR 'minor (person)'/exp OR 'puberty'/exp OR pediatric:ab,ti OR pediatrics:ab,ti OR paediatric:ab,ti OR paediatrics:ab,ti OR infant:ab,ti OR infants:ab,ti OR infantile:ab,ti OR baby:ab,ti OR babies:ab,ti OR preterm:ab,ti OR prematurity:ab,ti OR child:ab,ti OR children:ab,ti OR childhood:ab,ti OR toddler:ab,ti OR toddlers:ab,ti OR boy:ab,ti OR boys:ab,ti OR boyhood:ab,ti OR girl:ab,ti OR girls:ab,ti OR girlhood:ab,ti OR schoolchild:ab,ti OR schoolgirl:ab,ti OR schoolgirls:ab,ti OR schoolboy:ab,ti OR schoolboys:ab,ti OR 'school age':ab,ti OR preadolescent:ab,ti OR preadolescents:ab,ti OR preadolescence:ab,ti OR adolescent:ab,ti OR adolescents:ab,ti OR adolescence:ab,ti OR juvenile:ab,ti OR juveniles:ab,ti OR youth:ab,ti OR youths:ab,ti OR teenager:ab,ti OR teenagers:ab,ti OR teenaged:ab,ti OR teen:ab,ti OR teens:ab,ti OR pubescent:ab,ti OR puberty:ab,ti OR pubescence:ab,ti OR prepubescent:ab,ti OR prepubescence:ab,ti OR minors:ab,ti | 4606004 |
| 13 | 12 and 11 | 6481 |
| 14 | 13 NOT 'conference abstract'/it | 4428 |
| 15 | 17 AND [humans]/lim | 4106 |

Topic: How does experiencing adverse childhood experience changes children’s biologic reaction to stress and other biomarkers of health.

Searcher: SJK

Date: 4.27.2020

Database (including vendor/platform): Scopus

| Set |  | Results |
| --- | --- | --- |
| 1 | TITLE-ABS-KEY("toxic stress" OR ACEs OR ((child OR childhood OR children) W/3 (adverse or adversity or adversities))) | 69755 |
| 2 | TITLE-ABS-KEY (Biomarker or biomarkers or epigenomics OR epigenetic OR epigenetics OR interleukins OR interleukin OR TNF OR interferon OR interferons OR fibrinogen OR leukocyte OR leukocytes OR cytokine OR cytokines OR chemokines OR chemokine OR insulin OR telomere OR cortisol OR telomeres OR (Tumor W/1 Necrosis W/1 Factor W/1 alpha) OR (c W/1 reactive W/1 protein) OR CRP OR telomeres OR telomere or ((Biological OR bio OR immune OR serum OR plasma OR blood OR saliva OR urine OR laboratory OR viral OR clinical OR surrogate OR immunologic OR biochemical or molecular or genetic or inflammatory OR inflammation OR physiological OR cardiometabolic OR cardiovascular OR proinflammatory) W/2 (Marker OR markers OR endpoint OR levels OR level))) | 4308860 |
| 3 | TITLE-ABS-KEY (reaction or reactions or reacted or reactive or reacting or stress or stresses or stressor or stressors or stressed or adaptation or adaption or adapt or adapted or adapts or adapting or development or developing or developed or develop or develops or developmental or maladaptation or maladaption or maladapted or outcome or outcomes or impact or impacts or influence or influences or function or functions or functioning or functioned OR allostasis OR (allostatic W/1 load)) | 29958682 |
| 4 | TITLE-ABS-KEY (child or children or childhood or pediatric or pediatrics or paediatric or paediatrics or kid or kids or boy or girl or boys or girls or infant or infants or infancy or neonate or neonates or baby or babies or juvenile or juveniles or adolescent or adolescents or adolescence or teen or teens or teenager or teenagers or boyhood or girlhood or newborn or newborns) | 5404959 |
| 5 | 1 AND 2 AND 3 AND 4 | 1983 |

Searcher: SJK

Date: 4.28.2020

Database (including vendor/platform): PsycINFO

| Set |  | Results |
| --- | --- | --- |
| 1 | DE "Childhood Adversity" OR DE "Child Abuse" OR DE "Child Neglect" OR DE "Child Welfare" OR DE "Domestic Violence" OR DE "Emotional Abuse" OR DE "Physical Abuse" OR DE "Sexual Abuse" OR DE "Verbal Abuse" OR DE "Alcoholism" OR DE "Alcohol Abuse" OR DE "Suicide" OR DE "Divorce" OR DE "Racism" OR DE "Parental Absence" OR DE "Poverty" OR DE "Homeless" OR DE "Intimate Partner Violence" OR DE "Major Depression" | 305648 |
| 2 | (TI ACEs OR AB ACEs OR ((TI toxic OR AB toxic) N1 (TI stress OR AB stress)) OR ((TI childhood OR AB childhood) N1 (TI adversity OR AB adversity)) OR (TI "childhood trauma" OR AB "childhood trauma") OR ((TI adverse OR AB adverse) N1 (TI childhood OR AB childhood)) OR ((TI childhood OR AB childhood) N1 (TI adversities OR AB adversities)) OR ((TI Adverse OR AB Adverse OR TI adversity OR AB adversity OR TI adversities OR AB adversities) N3 (TI child OR AB child OR TI childhood OR AB childhood OR TI children OR AB children) N3 (TI exposure OR AB exposure OR TI exposures OR AB exposures OR TI exposed OR AB exposed OR TI event OR AB event OR TI events OR AB events OR TI experience OR AB experience OR TI experiences OR AB experiences OR TI experienced OR AB experienced OR TI experiencing OR AB experiencing))) | 9664 |
| 3 | 1 OR 2 | 310863 |
| 4 | DE "Biological Markers" | 12701 |
| 5 | (TI Biomarker OR AB Biomarker OR TI biomarkers OR AB biomarkers OR TI epigenomics OR AB epigenomics OR TI epigenetic OR AB epigenetic OR TI epigenetics OR AB epigenetics OR TI interleukins OR AB interleukins OR TI interleukin OR AB interleukin OR TI TNF OR AB TNF OR TI interferon OR AB interferon OR TI interferons OR AB interferons OR TI fibrinogen OR AB fibrinogen OR TI leukocyte OR AB leukocyte OR TI leukocytes OR AB leukocytes OR TI cytokine OR AB cytokine OR TI cytokines OR AB cytokines OR TI chemokines OR AB chemokines OR TI chemokine OR AB chemokine OR TI insulin OR AB insulin OR TI telomere OR AB telomere OR TI cortisol OR AB cortisol OR TI telomeres OR AB telomeres OR (TI Tumor OR AB Tumor N1 TI Necrosis OR AB Necrosis N1 TI Factor OR AB Factor N1 TI alpha OR AB alpha) OR (TI c OR AB c N1 TI reactive OR AB reactive N1 TI protein OR AB protein) OR TI CRP OR AB CRP OR TI telomeres OR AB telomeres OR TI telomere OR AB telomere OR ((TI Biological OR AB Biological OR TI bio OR AB bio OR TI immune OR AB immune OR TI serum OR AB serum OR TI plasma OR AB plasma OR TI blood OR AB blood OR TI saliva OR AB saliva OR TI urine OR AB urine OR TI laboratory OR AB laboratory OR TI viral OR AB viral OR TI clinical OR AB clinical OR TI surrogate OR AB surrogate OR TI immunologic OR AB immunologic OR TI biochemical OR AB biochemical OR TI molecular OR AB molecular OR TI genetic OR AB genetic OR TI inflammatory OR AB inflammatory OR TI inflammation OR AB inflammation OR TI physiological OR AB physiological OR TI cardiometabolic OR AB cardiometabolic OR TI cardiovascular OR AB cardiovascular OR TI proinflammatory OR AB proinflammatory) N2 (TI Marker OR AB Marker OR TI markers OR AB markers OR TI endpoint OR AB endpoint OR TI levels OR AB levels OR TI level OR AB level))) | 203021 |
| 6 | 4 or 5 | 206051 |
| 7 | 3 and 6 | 16179 |
| 8 | DE "Physiological Stress" OR DE "Post-Traumatic Stress" OR DE "Psychological Stress" OR DE "Stress Reactions" OR DE "Stress and Trauma Related Disorders" OR DE "Acute Stress Disorder" OR DE "Posttraumatic Stress Disorder" | 53237 |
| 9 | (TI reaction OR AB reaction OR TI reactions OR AB reactions OR TI reacted OR AB reacted OR TI reactive OR AB reactive OR TI reacting OR AB reacting OR TI stress OR AB stress OR TI stresses OR AB stresses OR TI stressor OR AB stressor OR TI stressors OR AB stressors OR TI stressed OR AB stressed OR TI adaptation OR AB adaptation OR TI adaption OR AB adaption OR TI adapt OR AB adapt OR TI adapted OR AB adapted OR TI adapts OR AB adapts OR TI adapting OR AB adapting OR TI development OR AB development OR TI developing OR AB developing OR TI developed OR AB developed OR TI develop OR AB develop OR TI develops OR AB develops OR TI developmental OR AB developmental OR TI maladaptation OR AB maladaptation OR TI maladaption OR AB maladaption OR TI maladapted OR AB maladapted OR TI outcome OR AB outcome OR TI outcomes OR AB outcomes OR TI impact OR AB impact OR TI impacts OR AB impacts OR TI influence OR AB influence OR TI influences OR AB influences OR TI function OR AB function OR TI functions OR AB functions OR TI functioning OR AB functioning OR TI functioned OR AB functioned OR TI allostasis OR AB allostasis OR (TI allostatic OR AB allostatic N1 TI load OR AB load)) | 2,326,627 |
| 10 | 8 or 9 | 2,330,372 |
| 11 | 7 and 10 | 9556 |
| 12 | MH "Adolescence+" OR MH "Child+" OR MH "Pediatrics+" OR MH "Minors (Legal)" OR MH "Puberty+" OR TI(Infant OR infants OR infancy OR newborn OR newborns OR neonatal OR neonate OR neonates OR baby OR babies OR preterm OR prematurity OR toddler OR toddlers OR boy OR boys OR boyhood OR girl OR girls OR girlhood OR kid OR kids OR child OR childhood OR children OR schoolchild OR schoolgirl OR schoolgirls OR schoolboy OR schoolboys OR "school age" OR "school aged" OR preadolescent OR preadolescents OR preadolescence OR adolescent OR adolescents OR adolescence OR juvenile OR juveniles OR youth OR youths OR teen OR teens OR teenager OR teenagers OR teenaged OR puberty OR pubescent OR pubescence OR prepubescent OR prepubescence OR pediatric OR pediatrics OR paediatric OR paediatrics OR minors) OR AB(Infant OR infants OR infancy OR newborn OR newborns OR neonatal OR neonate OR neonates OR baby OR babies OR preterm OR prematurity OR toddler OR toddlers OR boy OR boys OR boyhood OR girl OR girls OR girlhood OR kid OR kids OR child OR childhood OR children OR schoolchild OR schoolgirl OR schoolgirls OR schoolboy OR schoolboys OR "school age" OR "school aged" OR preadolescent OR preadolescents OR preadolescence OR adolescent OR adolescents OR adolescence OR juvenile OR juveniles OR youth OR youths OR teen OR teens OR teenager OR teenagers OR teenaged OR puberty OR pubescent OR pubescence OR prepubescent OR prepubescence OR pediatric OR pediatrics OR paediatric OR paediatrics OR minors) | 988388 |
| 13 | 11 AND 12 | 2213 |
|  | Limit to academic journals | 1910 |
