## Supplementary Table 2 for "Systematic Review and Meta-Analysis of the Effect of Adverse Childhood Experiences (ACEs) on Brain-Derived Neurotrophic Factor (BDNF) Levels"

**Supplementary Table 2: Characteristics of Studies Included within the Systematic Review**

| Author (year) | Study Population (Sample Size) | ACEs Measured, ACE Timing | BDNF Measurement Methodology, Measurement Timing | Impact of ACE exposure on BDNF levels |
| --- | --- | --- | --- | --- |
| Aksu, Unlu, Kardesler, Cakaloz, and Aybek (2018) | 9 comparator group; 22 ACE+ | Childhood sexual abuse, during childhood following ACE exposure | ELISA on serum, during childhood post ACE | Higher in ACE+ |
| Benedetti et al. (2017) | 5HTTLPR l/l (15),  5-HTTLPR l/s (18), 5-HTTLPR-s/s (7), 5HTTLPR-*/s (25) | Childhood abuse (sexual, physical, and/or emotional), neglect (physical and/or emotional), retrospectively in adulthood | ELISA on serum, during young, middle, and late adulthood (19-40+) | Lower in ACE+ |
| Bortoluzzi et al. (2014) | 50 comparator group; 40 ACE+ | Childhood abuse (sexual, physical, and/or emotional), neglect (physical and/or emotional), during childhood following ACE exposure | ELISA on serum, during childhood | Higher in ACE+ |
| Bücker et al. (2015) | 26 comparator group; 36 ACE+ | Childhood abuse (sexual, physical, and/or emotional) and neglect (physical and/or emotional), during childhood both during ACE exposure and following ACE exposure | ELISA on plasma, during childhood | Higher in ACE+ |
| Counotte et al. (2019) | 41 comparator group; 44 ACE+ | Childhood abuse (sexual, physical, and/or emotional), neglect (physical and/or emotional), retrospectively during adulthood | Luminex assay on serum, retrospectively in young and middle adulthood (19-40) | No significant difference in levels of BDNF |
| do Prado, Grassi-Oliveira, Daruy-Filho, Wieck, and Bauer (2017) | 27 comparator group; 30 ACE+ | Childhood Abuse (sexual, physical, and/or emotional abuse) and neglect (emotional and/or physical), during childhood following ACE exposure | ELISA on plasma, during childhood following ACE exposure | Lower in ACE+ |
| Druzhkova et al. (2019) | 613 comparator group; 822 ACE+ | Childhood abuse (sexual, physical, and/or emotional), neglect (physical and/or emotional), retrospectively in adulthood | ELISA on serum, during young, middle, and late adulthood (19-40+) | Lower in the ACE+ |
| Grassi-Oliveira, Stein, Lopes, Teixeira, and Bauer (2008) | 32 comparator group; 17 ACE+ | Childhood neglect (emotional and/or physical), retrospectively in adulthood | ELISA on plasma, during young, middle, and late adulthood (19-40+) | Lower in ACE+ |
| Hauck et al. (2010) | 34 comparator group; 13 ACE+ | Not explicitly reported (defined as remote trauma), retrospectively in adulthood | ELISA on serum, retrospectively in young and middle adulthood (19-40+) | Higher in ACE+ |
| Kauer‐Sant'Anna et al. (2007) | 85 comparator group; 78 ACE+ | Childhood abuse (sexual and physical), retrospectively in adulthood | ELISA on serum, retrospectively in middle and late adulthood (24-40+) | Lower in ACE+ |
| Kavurma et al. (2017) | 35 comparator group; 70 ACE+ | Childhood abuse (sexual, physical, and/or emotional), neglect (physical and/or emotional), during childhood following ACE exposure | ELISA on serum, during childhood | Lower in ACE+ |
| Mansur et al. (2016) | 28 comparator group; 467 ACE+ | Peer victimization/Bullying, Socioeconomic disadvantage, during childhood while ACE exposure ongoing | ELISA on serum, during childhood | Higher in the ACE+ |
| Palmos et al. (2019) | 301 comparator group; 163 ACE+ | Childhood abuse (sexual, physical, and/or emotional) and neglect (physical and/or emotional), retrospectively during adulthood | ELISA on serum, during middle and late adulthood (25-40+) | No significant difference between groups |
| Sharma, Graham, Rohde, and Ceballos (2017) | 22 comparator group;  46 ACE+ | Family history of substance abuse (alcohol use disorder), retrospectively in adulthood | ELISA on serum, retrospectively during young and middle adulthood (19-40+) | Lower in ACE+ |
| Simsek, Uysal, Kaplan, Yuksel, and Aktas (2015) | 28 comparator group; 27 ACE+ | Childhood sexual abuse, during childhood following ACE exposure | ELISA on blood, during childhood following ACE exposure | Lower in ACE+ |
| Snijders et al. (2017) | 80 comparator group; 146 ACE+ | Exposure to parent with bipolar disorder, during childhood while ACE exposure ongoing | ELISA on Serum, during childhood | Lower in ACE+ |
| Sordi et al. (2019) | 11 comparator group; 22 ACE+ | Childhood abuse (sexual, physical, and/or emotional), retrospectively in adulthood | ELISA on blood, during young and middle adulthood (19-40) | Higher in ACE+ group |
| Theleritis et al. (2014) | 152 comparator group; 95 ACE+ | Childhood abuse (sexual and/or physical) and parental death, retrospectively in adulthood | ELISA on plasma, retrospectively in young, middle, and late Adulthood (19-40+) | Lower in ACE+ |
| Trajkovska, Vinberg, Aznar, Knudsen, and Kessing (2008) | 35 comparator group; 26 ACE+ | Parental mental health, retrospectively in adulthood | ELISA on blood, retrospectively during early, middle, and late adulthood (19-40+) | Lower in ACE+ |
| van der Meij, Comijs, Dols, Janzing, and Voshaar (2014) | 231 comparator group; 108 ACE+ | Sexual abuse, retrospectively in adulthood | ELISA on serum, during late adulthood (40+) | Higher in ACE+ |
| Viola et al. (2014) | 82 comparator group; 22 ACE+ | Childhood sexual abuse, retrospectively in adulthood | ELISA on plasma, during young, middle, and late adulthood (19-40+) | Higher in ACE+ |
| Watt, Ceballos, Kim, Pan, and Sharma (2020) | 30 comparator group; 63 ACE+ | Parental mental health, parental substance abuse, parental incarceration, and exposure to intimate partner violence, retrospectively in adulthood | ELISA on serum, retrospectively in young Adulthood (19-24) | Lower in ACE+ |

Works Cited

Aksu, S., Unlu, G., Kardesler, A. C., Cakaloz, B., & Aybek, H. (2018). Altered levels of brain-derived neurotrophic factor, proBDNF and tissue plasminogen activator in children with posttraumatic stress disorder. *Psychiatry Res, 268*, 478-483.

Benedetti, F., Ambree, O., Locatelli, C., Lorenzi, C., Poletti, S., Colombo, C., & Arolt, V. (2017). The effect of childhood trauma on serum BDNF in bipolar depression is modulated by the serotonin promoter genotype. *Neurosci Lett, 656*, 177-181.

Bortoluzzi, A., Salum, G. A., Blaya, C., Silveira, P. P., Grassi-Oliveira, R., da Rosa, E. D., . . . Schuch, I. (2014). Mineralocorticoid receptor genotype moderates the association between physical neglect and serum BDNF. *Journal of psychiatric research, 59*, 8-13.

Bücker, J., Fries, G., Kapczinski, F., Post, R., Yatham, L., Vianna, P., . . . Aguiar, B. (2015). Brain‐derived neurotrophic factor and inflammatory markers in school‐aged children with early trauma. *Acta Psychiatrica Scandinavica, 131*(5), 360-368.

Counotte, J., Bergink, V., Pot-Kolder, R., Drexhage, H. A., Hoek, H. W., & Veling, W. (2019). Inflammatory cytokines and growth factors were not associated with psychosis liability or childhood trauma. *PLoS One, 14*(7), e0219139.

do Prado, C. H., Grassi-Oliveira, R., Daruy-Filho, L., Wieck, A., & Bauer, M. E. (2017). Evidence for immune activation and resistance to glucocorticoids following childhood maltreatment in adolescents without psychopathology. *Neuropsychopharmacology, 42*(11), 2272-2282.

Druzhkova, T., Pochigaeva, K., Yakovlev, A., Gersamia, A., Guekht, A., & Gulyaeva, N. (2019). Effects of Childhood Trauma on the Biological Correlates of Stress in Men and Women with Borderline Mental Disorders. *Neuroscience and Behavioral Physiology, 49*(7), 916-920.

Grassi-Oliveira, R., Stein, L. M., Lopes, R. P., Teixeira, A. L., & Bauer, M. E. (2008). Low plasma brain-derived neurotrophic factor and childhood physical neglect are associated with verbal memory impairment in major depression—a preliminary report. *Biological psychiatry, 64*(4), 281-285.

Hauck, S., Kapczinski, F., Roesler, R., de Moura Silveira Jr, É., Magalhães, P. V., Kruel, L. R. P., . . . Ceitlin, L. H. F. (2010). Serum brain-derived neurotrophic factor in patients with trauma psychopathology. *Progress in Neuro-Psychopharmacology and Biological Psychiatry, 34*(3), 459-462.

Kauer‐Sant'Anna, M., Tramontina, J., Andreazza, A. C., Cereser, K., Costa, S. d., Santin, A., . . . Kapczinski, F. (2007). Traumatic life events in bipolar disorder: impact on BDNF levels and psychopathology. *Bipolar Disorders, 9*, 128-135.

Kavurma, C., Tas, F. V., Demirgoren, B. S., Demirci, F., Akan, P., Eyuboglu, D., & Guvenir, T. (2017). Do serum BDNF levels vary in self-harm behavior among adolescents and are they correlated with traumatic experiences? *Psychiatry Res, 258*, 130-135.

Mansur, R. B., Cunha, G. R., Asevedo, E., Zugman, A., Zeni-Graiff, M., Rios, A. C., . . . Gadelha, A. (2016). Socioeconomic disadvantage moderates the association between peripheral biomarkers and childhood psychopathology. *PLoS One, 11*(8), e0160455.

Palmos, A. B., Watson, S., Hughes, T., Finkelmeyer, A., McAllister-Williams, R. H., Ferrier, N., . . . Strawbridge, R. (2019). Associations between childhood maltreatment and inflammatory markers. *BJPsych open, 5*(1).

Sharma, S., Graham, R., Rohde, R., & Ceballos, N. A. (2017). Stress-induced change in serum BDNF is related to quantitative family history of alcohol use disorder and age at first alcohol use. *Pharmacology Biochemistry and Behavior, 153*, 12-17.

Simsek, S., Uysal, C., Kaplan, I., Yuksel, T., & Aktas, H. (2015). BDNF and cortisol levels in children with or without post-traumatic stress disorder after sustaining sexual abuse. *Psychoneuroendocrinology, 56*, 45-51.

Snijders, G., Mesman, E., de Wit, H., Wijkhuijs, A., Nolen, W., Drexhage, H., & Hillegers, M. (2017). Immune dysregulation in offspring of a bipolar parent. Altered serum levels of immune growth factors at adolescent age. *Brain, behavior, and immunity, 64*, 116-123.

Sordi, A. O., von Diemen, L., Kessler, F. H., Schuch, S., Ornell, F., Kapczinski, F., . . . Salum, G. A. (2019). Effects of childhood trauma on BDNF and TBARS during crack-cocaine withdrawal. *Brazilian Journal of Psychiatry, 42*, 214-217.

Theleritis, C., Fisher, H. L., Shäfer, I., Winters, L., Stahl, D., Morgan, C., . . . Vitoratou, S. (2014). Brain derived neurotropic factor (BDNF) is associated with childhood abuse but not cognitive domains in first episode psychosis. *Schizophr Res, 159*(1), 56-61.

Trajkovska, V., Vinberg, M., Aznar, S., Knudsen, G. M., & Kessing, L. V. (2008). Whole blood BDNF levels in healthy twins discordant for affective disorder: association to life events and neuroticism. *Journal of affective disorders, 108*(1-2), 165-169.

van der Meij, A., Comijs, H. C., Dols, A., Janzing, J. G., & Voshaar, R. C. O. (2014). BDNF in late-life depression: effect of SSRI usage and interaction with childhood abuse. *Psychoneuroendocrinology, 43*, 81-89.

Viola, T. W., Tractenberg, S. G., Levandowski, M. L., Pezzi, J. C., Bauer, M. E., Teixeira, A. L., & Grassi-Oliveira, R. (2014). Neurotrophic factors in women with crack cocaine dependence during early abstinence: the role of early life stress. *Journal of Psychiatry and Neuroscience, 39*(3), 206-214.

Watt, T., Ceballos, N., Kim, S., Pan, X., & Sharma, S. (2020). The unique nature of depression and anxiety among college students with adverse childhood experiences. *Journal of child & adolescent trauma, 13*(2), 163-172.
